# VNtyper 2 enables open-access short-read genotyping of MUC1 VNTR variants in ADTKD at high-speed

**DOI:** 10.64898/2026.05.27.26352937

**Authors:** Bernt Popp, Hassan Saei, Omri Teltsh, Václav Janoušek, Anna Přistoupilová, Alena Vrbacká, Hana Hartmannová, Kendrah Kidd, Johannes Helmuth, Anthony J Bleyer, Michael Wiesener, Kathrin Fausch, Colm Rowan, Elhussein A. E. Elhassan, Michelle Clince, Gianpiero L. Cavalleri, Maurus Locher, Kai-Uwe Eckardt, Paulina Richter-Pechanska, ADTKD-Net Consortium, Stanislav Kmoch, Corinne Antignac, Peter Conlon, Guillaume Dorval, Martina Živná, Jan Halbritter

## Abstract

**Introduction:** ADTKD-*MUC1* is one of the major entities of ADTKD caused by frameshift variants in the *MUC1* VNTR that standard short-read sequencing fails to detect. Existing 59dupC-targeted probe-extension assays do not allow for broad screening and miss atypical non-dupC variants and 59dupC in repeat units with altered primer-binding sequence. Recently, VNtyper, a Kestrel-based genotyping pipeline with optional code-adVNTR cross-validation for *MUC1* VNTR genotyping from short-read data circumvented this limitation, but needed further development for easy access and rapid sample processing.

**Methods:** VNtyper 2 is a modular, production-grade tool for *MUC1* VNTR genotyping from short-read data, with a companion web platform, VNtyper-Online (https://vntyper.org), for freely available browser-based analysis with short turnaround and without local bioinformatics infrastructure. We validated VNtyper 2 on 2000 simulated samples and 142 clinical exomes with independently confirmed genotypes.

**Results:** In simulation, VNtyper 2 detected the canonical 59dupC variant with 97.2% sensitivity and 100.0% specificity. Reference-standard validation on 142 samples yielded 90.6% sensitivity and 98.2% specificity overall, with cohort-dependent performance across the Twist Exome v2 French-German cohort (98.0% sensitivity, 87.5% specificity) and the KAPA HyperExome V2 Czech-US cohort (79.4% sensitivity, 100.0% specificity). Screening of 3583 exomes and targeted panels from international CKD referral programmes identified 53 positive individuals, including 9 with atypical non-dupC frameshift variants that targeted assays would miss. In unselected CKD cohorts, a descriptive random-effects summary estimated a detection rate of 1.4% (95% CI 0.7%).

**Conclusions:** VNtyper 2 and VNtyper-Online are open-source tools for *MUC1* VNTR genotyping from short-read data. They support screening and triage in locally validated workflows when VNTR coverage is adequate, with orthogonal testing to confirm positive calls and to follow up negative results under strong clinical suspicion. Fast and freely accessible, they make *MUC1* genotyping widely available. For routine diagnostics, we propose an expert-informed two-pathway workflow developed through European ADTKD-Net consortium consensus.

**Lay Summary:** Some families inherit a kidney disease caused by changes in a gene called *MUC1*. These changes sit in a long, repetitive part of the gene that routine genetic tests cannot read, so many affected people remain undiagnosed. We developed VNtyper 2, a free computer program with a website (https://vntyper.org), that searches existing genetic test data for these changes within minutes, without new laboratory work. In 142 patient samples with known results, it identified 90.6% of those carrying a change. Searching 3583 genetic tests from kidney clinics in four European countries, it found 53 people with likely disease-causing changes, including 9 that common targeted tests would miss. VNtyper 2 lets laboratories diagnose this disease from data they already have, with a second test to confirm each positive result.

## Introduction

Autosomal dominant tubulointerstitial kidney disease caused by *MUC1* variants (ADTKD-*MUC1*) is one of the major entities of ADTKD, a hereditary kidney disorder marked by progressive chronic kidney disease (CKD), bland urinary sediment, and absent or minimal proteinuria.^1,2^ Clinical presentations are nonspecific and typically involve slowly declining kidney function that progresses to kidney failure between the third and seventh decades of life.^1^ Compared with ADTKD-*UMOD*, gout is less frequent and occurs later in ADTKD-*MUC1*; when present, hyperuricaemia and gout are considered secondary to progressive CKD rather than primary manifestations.^1,3^ Although the genes *MUC1* and *UMOD* account for the vast majority of ADTKD cases,^3^ ADTKD-*MUC1* is very likely underdiagnosed, because it is clinically nonspecific and genetic testing for it is hard to access.

The disease is driven by frameshift variants within the guanine-cytosine (GC)-rich variable number tandem repeat (VNTR) region of *MUC1*, which encodes mucin-1.^2,4,5^ These variants produce the aberrant frameshifted protein MUC1-fs.

Despite the considerable impact of ADTKD-*MUC1* on kidney outcomes, disease-specific therapies remain elusive. Current management follows supportive CKD care, including blood pressure management, treatment of CKD complications, and timely planning for kidney replacement therapy.^1,6^ The toxic gain-of-function mechanism of MUC1-fs has prompted development of targeted therapies aimed at clearing the aberrant protein.^2^ Timely diagnosis also has immediate clinical relevance: it can end a diagnostic odyssey, inform family planning and cascade testing, and guide surveillance.

Reliable diagnostic methods are therefore critical, yet standard short-read sequencing often fails to detect *MUC1* frameshift variants because of the VNTR’s high GC content and repetitive nature.^5,7^ We use 59dupC for the recurrent canonical cytosine duplication in the *MUC1* VNTR, also reported as 27dupC or c.428dupC, and term all other pathogenic VNTR frameshifts atypical non-dupC variants.^8–10^ These limitations prompted two classes of specialized methods: 59dupC-targeted assays, including SNaPshot minisequencing and mass spectrometry-based formats, and long-read sequencing, with PacBio single-molecule real-time (SMRT) sequencing as the established reference and Oxford Nanopore as an emerging alternative, which can span the entire VNTR and precisely localize variants.^1,11–14^ Although accurate, these methods are technically demanding, costly and available in very few specialized laboratories, which precludes large-scale use and standard diagnostic workflows.^13^ The diagnostic gap is substantial: in a Japanese cohort of 48 clinicopathologically suspected ADTKD patients, standard short-read sequencing detected only one *MUC1* variant, whereas long-read sequencing of the short-read-negative patients identified eight more.^15^ Immunodetection of MUC1-fs in kidney biopsies or urinary cells is a non-genetic alternative,^16^ but it requires prior clinical suspicion, biopsy-based testing also requires tissue, and neither is routine in diagnostic workflows.^5,17^

These laboratory techniques are complemented by bioinformatic methods. We previously developed or co-developed short-read tools for *MUC1* VNTR genotyping (Figure 1a).^18,19^ VNtyper identifies canonical 59dupC and atypical non-dupC frameshift variants and has shown high concordance with established assays.^18,20^ Subsequent French, Danish, and Japanese studies applied VNtyper to further cohorts, supporting its analytical validity.^8,20–22^ Here, we present VNtyper 2, a successor that keeps this Kestrel-based genotyping concept with faster runtime, reproducible software and graded reporting, and VNtyper-Online for browser-based use (Figure 1a,b; Supplementary Methods S1–S2). We used both to screen international CKD cohorts and propose a testing workflow for ADTKD-*MUC1*.

**Figure 1:**
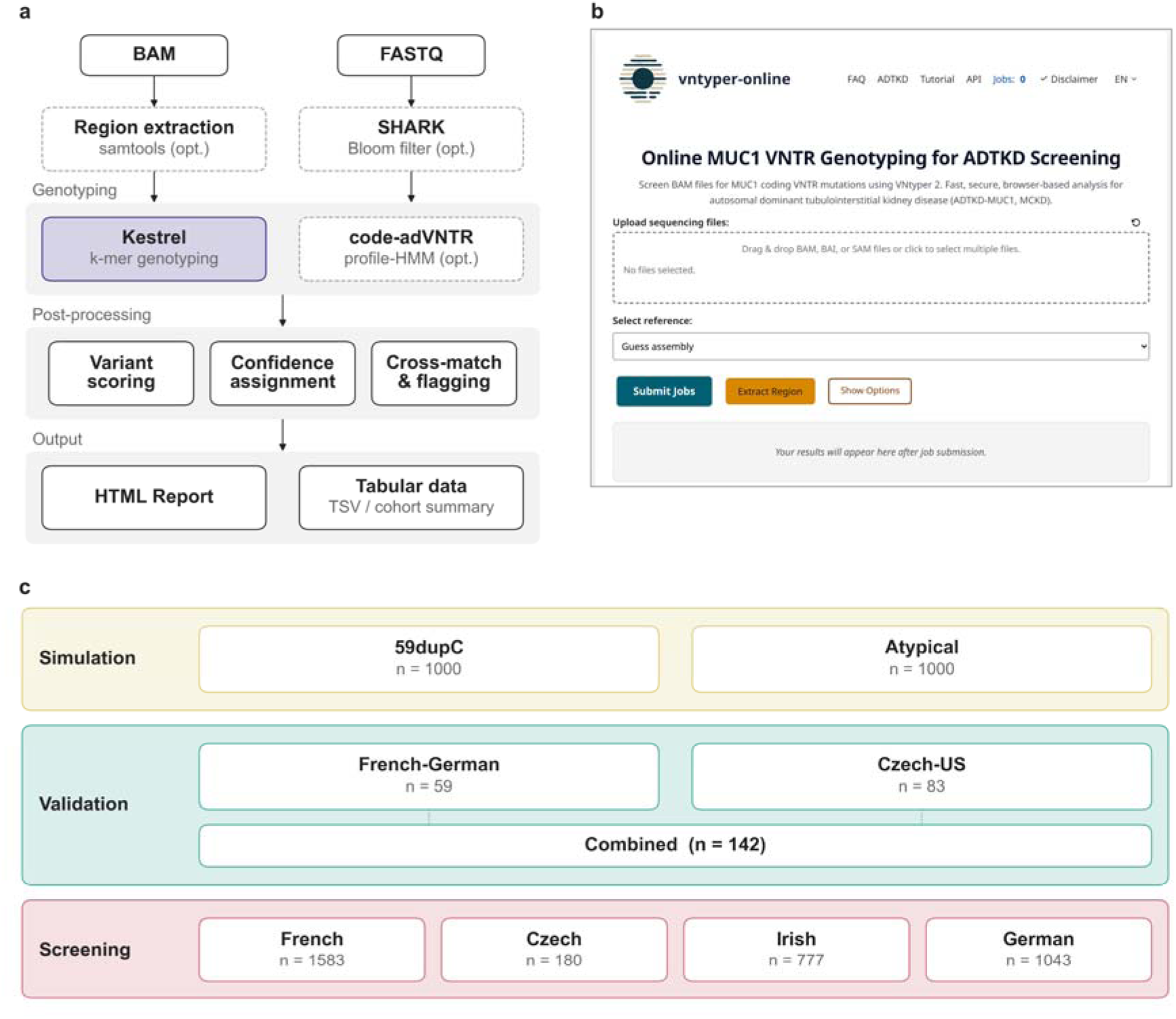
VNtyper 2 overview. (**a**) Pipeline architecture. BAM or FASTQ files undergo optional preprocessing before Kestrel (k-mer genotyping) and, optionally, code-adVNTR (profile-HMM). Post-processing assigns confidence labels and flags concordant calls. (**b**) VNtyper-Online (https://vntyper.org): browser-based *MUC1* screening with client-side region extraction via WebAssembly. (**c**) Study design. Simulated data (N = 2000; 59dupC and atypical non-dupC variants), gold standard with SNaPshot/PacBio-confirmed genotypes (N = 142), and screening cohorts from French (Renome, N = 1583), Czech (N = 180), Irish (N = 777), and German (N = 1043) CKD programmes (total screened N = 3583).

## Methods

### Cohorts examined

We assembled validation and screening cohorts from international centres (Figure 1c; Table 1). For validation, we used two independent gold standard cohorts totalling 142 samples with orthogonally confirmed genotypes: the French-German cohort (59 samples; 40 French samples previously confirmed by SNaPshot and PacBio long-read sequencing, and 19 German samples previously confirmed by SNaPshot from published families)^12,13^ and the Czech-US cohort (83 samples captured with KAPA HyperExome V2 and confirmed by PacBio; Supplementary Methods S5). Screening cohorts comprised French targeted panels (1583 samples from the Renome study at the Imagine Institute), Czech exomes (180 samples), Irish exomes (777 samples) recruited as part of the Irish Kidney Gene Project, and German exomes (1043 samples from the Charité Center for Rare Kidney Diseases (CeRKiD) registry) from CKD referral programmes.

**Table 1.** Cohort summary.

| Cohort | Category | N | Capture kit | Final calls (HP/LP; candidates) | Sens. / Det. rate (95% CI) | Spec. (95% CI) |
|---|---|---|---|---|---|---|
| ***A. Simulation & validation*** |  |  |  |  |  |  |
| MucOneUp dupC | Simulation | 1000 | Simulated | 500+ / 500– | 97.2% (95.4%–98.3%) | 100.0% (99.2%–100.0%) |
| MucOneUp atypical | Simulation | 1000 | Simulated | 500+ / 500– | 82.0% (78.4%–85.1%) | 100.0% (99.2%–100.0%) |
| French-German | Gold standard | 59 | Twist v2 | 51+ / 8– | 98.0% (89.7%–99.7%) | 87.5% (52.9%–97.8%) |
| Czech-US | Gold standard | 83 | KAPA HEv2 | 34+ / 49– | 79.4% (63.2%–89.7%) | 100.0% (92.7%–100.0%) |
| **Combined** | **Gold standard** | **142** |  | **85+ / 57–** | **90.6% (82.5%–95.2%)** | **98.2% (90.7%–99.7%)** |
| ***B. Screening*** |  |  |  |  |  |  |
| French | Unselected CKD | 1583 | Twist panel | 18 (18/0); +2 cand. | 1.14% (0.7%–1.8%) | – |
| Irish | Unselected CKD | 777 | Twist + CeGaT | 15 (15/0) | 1.93% (1.2%–3.2%) | – |
| Czech | Suspected ADTKD | 180 | KAPA HEv2 | 6 (4/2) | 3.33% (1.5%–7.1%) | – |
| German | Unselected CKD | 1043 | Twist v2 | 14 (9/5) | 1.34% (0.8%–2.2%) | – |

Section A shows simulation benchmarks and gold standard validation with test performance metrics. Section B shows retrospective screening cohorts with final detection yields and candidate calls. Sens., sensitivity; Det. rate, detection rate; Spec., specificity; HP, High Precision (combines High Precision and High Precision\*); LP, Low Precision; cand., candidate calls not counted as final positives; KAPA HEv2, KAPA HyperExome V2; CKD, chronic kidney disease; ADTKD, autosomal dominant tubulointerstitial kidney disease. Detection rates use 95% Wilson score confidence intervals. Sensitivity and specificity are reported only for cohorts with independent ground truth.

All cohorts underwent Illumina short-read sequencing, with mean depth over the unique sequence flanking the VNTR of approximately 133× in the Czech exomes, 187× in the Irish exomes, 200× in the German CeRKiD exomes and 593× in the French targeted panels (Supplementary Table S2; Supplementary Methods S4). All analyses used VNtyper 2.0.47.

### VNtyper 2 pipeline

We substantially extended VNtyper into a modular, production-grade pipeline with comprehensive test coverage and continuous integration (Supplementary Methods S1). Genotyping relies on Kestrel, a k-mer frequency-based variant caller that operates without alignment to a linear reference.^18,23^ We enlarged its state space and added a haplotype-count support metric. For raw sequencing reads (FASTQ), an optional upstream module re-implements the SHARK-based read filtering of the SharkVNTyper workflow, published without publicly available code, and passes only *MUC1*-relevant reads to Kestrel^8,24^ (Supplementary Methods S6).

A configurable flagging system annotates known false-positive patterns and low-depth variants in conserved motifs, and a cross-matching module compares Kestrel calls with code-adVNTR.^19^ Our maintained code-adVNTR fork runs in seconds per exome, so VNtyper-Online runs both callers by default. The pipeline accepts BAM, CRAM and FASTQ input and generates interactive reports; installation, containers and Snakemake batch processing are described in Supplementary Methods S1.

### Reference build, coordinates, and coverage

VNtyper 2 accepts alignments to GRCh37 (hg19) or GRCh38 (hg38) and detects the build and the chromosome naming (UCSC, Ensembl or RefSeq) from the file header. Reads are extracted from chr1:155,158,000-155,163,000 (GRCh37) or chr1:155,184,000-155,194,000 (GRCh38). Coverage is reported over a window spanning the repeat array (chr1:155,160,500-155,162,000 or chr1:155,188,000-155,192,500) and over 190 bp of unique flank on each side of the array (chr1:155,161,000-155,161,810 or chr1:155,188,530-155,192,010; 1-based, inclusive). Flank depth is comparable between builds; window depth is not, because GRCh37 holds fewer repeat units.

Each report gives a coverage verdict: PASS at a mean window coverage of at least 100×, FAIL below 50× and REDUCED in between on GRCh38 (290× and 145× on GRCh37). A sample confidence grade combines the call, flags, agreement between the two callers and the coverage verdict (Supplementary Methods S1).

### VNtyper-Online

We developed a web platform at https://vntyper.org (Figure 1b) that processes sequencing files in the user’s browser using biowasm (https://biowasm.com). Only the extracted *MUC1* region is transmitted to the backend for Kestrel genotyping; the full sequencing file never leaves the browser. The frontend and backend are open source and can be deployed locally using containerised infrastructure (Supplementary Methods S2).

### Simulation and benchmarking

We benchmarked VNtyper 2 on a prespecified simulation generated with MucOneUp 0.53.0,^25^ with seeds held out from all development runs (Supplementary Methods S3). The natural-length simulation comprised 500 matched wild-type and mutated pairs for 59dupC and 500 for ten atypical non-dupC frameshifts, with alleles within the range of PacBio-resolved lengths (35 to 105 repeats)^10^ and a coverage titration from 75% to 6.25%. A separate long-allele simulation (200 pairs) placed the variant on an allele of 105 to 140 repeats and is reported separately. Both experiments ran Kestrel and code-adVNTR.

### Statistical analysis

Sensitivity was defined as true positives (TP) divided by true positives plus false negatives (FN), and specificity as true negatives (TN) divided by true negatives plus false positives (FP). Ground truth was the MucOneUp label for simulated data and SNaPshot minisequencing or PacBio long-read sequencing for clinical samples. We calculated 95% confidence intervals with the Wilson score method. To compare versions, we ran the original VNtyper 1.3.0 (Kestrel only) on the same reference-standard alignments with identical counting rules (Supplementary Methods S7). We stratified performance by cohort, capture kit and depth with Firth-penalised logistic regression (Supplementary Methods S10), swept the depth-score threshold (Supplementary Methods S8), and downsampled confirmed positive exomes to model coverage-dependent sensitivity (Supplementary Methods S9). Detection rates of the three unselected screening cohorts were pooled with a logit-scale random-effects model; with three cohorts, its confidence interval and heterogeneity statistics are descriptive (Supplementary Methods S6). All analyses were performed in Python.

### Study approval

All cohorts were collected under local institutional review board approvals, and written informed consent was obtained from each participant. Specific ethics committee names and approval details are provided in Supplementary Methods S4.

## Results

### VNtyper-Online enables browser-based *MUC1* genotyping

We developed VNtyper-Online (https://vntyper.org), a browser-based platform that allows analysis of *MUC1* VNTR variants (Figure 1b). Users upload BAM/BAI or SAM files (e.g. exported from IGV) through a drag-and-drop interface, and the platform automatically detects the reference genome assembly from file headers, supporting six naming conventions across hg19, hg38, GRCh37 and GRCh38. A WebAssembly-compiled samtools instance running in the browser extracts only the *MUC1* locus; the full sequencing file never leaves the user’s machine. Only this extracted region is transmitted to the server for Kestrel genotyping (Supplementary Methods S2). On identical *MUC1* region slices from 89 exomes, the public instance and a local run of the same version gave identical results, with a median of 18.8 s per sample online and 4.3 s locally (Supplementary Methods S2).

The platform supports both individual case analysis and larger cohort analyses. An optional cohort mode groups multiple samples under a shared alias protected by a passphrase, enabling batch submission and joint download of results with email notification upon completion. Code-adVNTR runs by default for cross-validation, and normal mode includes unmapped reads for more thorough variant detection without materially increasing runtime for standard exomes. An interactive tutorial and built-in FAQ guide new users through the interface.

The public VNtyper-Online instance was designed as a privacy-conscious demonstration and research service. All communication uses HTTPS/TLS, the deployment had an independently verified A+ security rating at assessment, uploaded regional data are deleted immediately after analysis, and results are purged after seven days.

As of 6 September 2026, the public instance had processed 4312 analyses since its launch on 21 November 2024, indicating international uptake by users including centres without local VNtyper installation or dedicated bioinformatics support. This figure reflects an aggregate analysis count only; the platform does not retain or expose per-analysis results to the operators.

### VNtyper 2 achieves high sensitivity for both canonical and atypical variants

We assessed VNtyper 2 on a prespecified, held-out simulation generated with the MucOneUp simulator we developed^25^ (Figure 2a,b). For the canonical 59dupC variant, Kestrel detected 486 of 500 positive samples (sensitivity 97.2%, 95% CI 95.4%–98.3%) with 100.0% specificity (95% CI 99.2%–100.0%). We then tested ten atypical non-dupC frameshift types (500 pairs). Aggregate sensitivity was 82.0% (95% CI 78.4%–85.1%), with per-type sensitivity ranging from 46.0% (insG_pos54) to 98.0% (insCCCC). Specificity remained 100.0% across all variant types. Coverage titration showed that 59dupC sensitivity remained at 80% or more down to 25% of full coverage (Supplementary Figure S2). Longer mutated alleles were detected less often (odds ratio 0.48 per 10 repeats for 59dupC; Supplementary Table S1), and counting a call by either Kestrel or code-adVNTR raised atypical sensitivity to 93.8% with specificity unchanged. With the variant on an allele of 105 to 140 repeats, sensitivity was 58.0% for 59dupC and 29.0% for atypical variants, again without false positives (Supplementary Figure S4).

**Figure 2:**
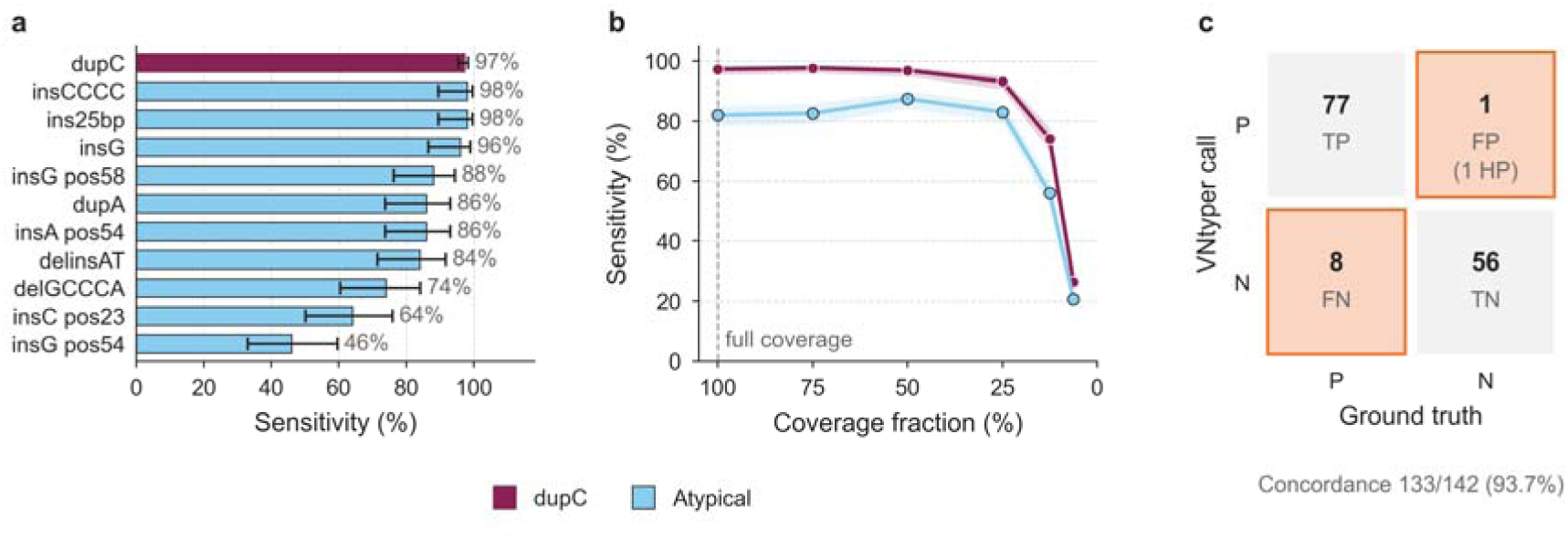
Simulation performance and gold standard validation. (a) Per-variant Kestrel sensitivity of VNtyper 2 in the prespecified held-out MucOneUp simulation (natural-length simulation, alleles of 35 to 105 repeats). The canonical 59dupC variant (wine; 500 pairs) was detected in 97.2% of cases, and ten atypical non-dupC frameshifts (light blue; 50 pairs each) ranged from 46.0% (insG_pos54) to 98.0% (insCCCC). Error bars indicate Wilson 95% confidence intervals. Specificity was 100.0% for every variant type (no false positives among 1000 wild-type samples). (**b**) Sensitivity as a function of VNTR-coverage fraction. 59dupC sensitivity stayed at 80% or more down to 25% of full coverage before declining sharply; specificity was 100.0% at every fraction. Shaded bands indicate 95% confidence intervals. (**c**) Combined gold standard concordance matrix. P indicates positive and N indicates negative. VNtyper 2 calls were compared against PacBio/SNaPshot-confirmed genotypes across 142 exome samples from two validation cohorts with different ascertainment and reference-standard composition (French-German and Czech-US). Overall concordance was 93.7% with 77 true positives and 8 false negatives. See Supplementary Figures S1–S4 and Supplementary Table S1 for length effects, the variant-by-coverage heatmap, false negatives, and long alleles (long-allele simulation).

The two clinical cohorts (Figure 2c) differed in ascertainment: the French-German set (59 Twist Exome v2 samples; 51 positive, 8 negative) was enriched for SNaPshot-confirmed 59dupC cases, whereas the Czech-US set (83 KAPA HyperExome V2 samples; 34 positive, 49 negative) consisted of unresolved high-suspicion referrals resolved by PacBio. In the French-German cohort, VNtyper 2 detected 50 of 51 positives (sensitivity 98.0%, 95% CI 89.7%–99.7%) with one discordant call (specificity 87.5%, 95% CI 52.9%–97.8%). This sample (FR-GS-006) was called positive in two independent sequencing runs, by code-adVNTR and by the original VNtyper, but was negative by PacBio with an uninterpretable SNaPshot result; we count it as a false positive (Supplementary Methods S7). In the Czech-US cohort, sensitivity was 79.4% (27/34; 95% CI 63.2%–89.7%) with 100.0% specificity. Across both cohorts (142 samples), sensitivity was 90.6% (95% CI 82.5%–95.2%) and specificity 98.2% (95% CI 90.7%–99.7%), with 66 of 70 59dupC and 11 of 15 atypical carriers detected (Supplementary Table S3). Predictive values in the per-cohort confusion matrices (Supplementary Figure S6) describe these enriched cohorts, not unselected testing.

Sensitivity was 98.0% with Twist Exome v2 and 79.4% with KAPA HyperExome V2. The difference persisted after adjustment for flank depth, although kit coincides with cohort, centre and variant spectrum (Supplementary Methods S10; Table S3). Of 4 carriers missed on KAPA data and resequenced from the same DNA with Twist Exome v2, VNtyper 2 called 3 (Supplementary Methods S11; Table S12). Supplementary Tables S8 and S10 list every false-negative and discordant case with its reference standard.

The shipped depth-score threshold sat on a stable plateau (pooled area under the receiver operating characteristic curve 0.978). Lowering it would have recovered few false negatives at the cost of false positives among the French negatives, so we kept it and provide local calibration for other assays (Supplementary Methods S8; Figure S8; Table S9).

On the same 142 reference-standard exomes, VNtyper 2 matched the sensitivity of VNtyper 1.3.0, filtered the one 4-bp insertion artifact that VNtyper 1.3.0 reported as a call (specificity 98.2% versus 96.5%), and completed a full run including code-adVNTR 107-fold faster (Supplementary Methods S7; Tables S6 and S7).

### Retrospective screening of CKD cohorts detects ADTKD-*MUC1* positives

We screened 3583 exomes and targeted panels from four international CKD referral programmes (Figure 3a). Among the unselected cohorts, the French cohort (n = 1583 targeted panels) yielded 18 final positives (1.14%); two further low-precision (LP) 59dupC calls with low alternate-read support remain unconfirmed candidates. The Irish cohort (n = 777 exomes) yielded 15 positives (1.93%; 15 high precision), and the German CeRKiD cohort (n = 1043 exomes) 14 (1.34%; 9 high precision and 5 low precision). The pooled detection rate of these three cohorts was 1.4% (95% CI 0.7%–2.8%), including atypical variants. With the 59dupC-focused SharkVNTyper and no sample overlap with our study, Bensouna et al. reported a similar rate of 1.4% overall (63/4653) and 1.5% in their prospective CKD screening subset.^8^ The Czech cohort (n = 180 samples), which included individuals with prior clinical suspicion of ADTKD, had the expected higher rate of 3.33% (6 positives), consistent with the 5.7% reported by Granhoj et al. in Danish patients with kidney failure of undetermined aetiology at age 50 or younger^22^ (Figure 3a). Most final positive calls were high precision (Figure 3b,c).

**Figure 3:**
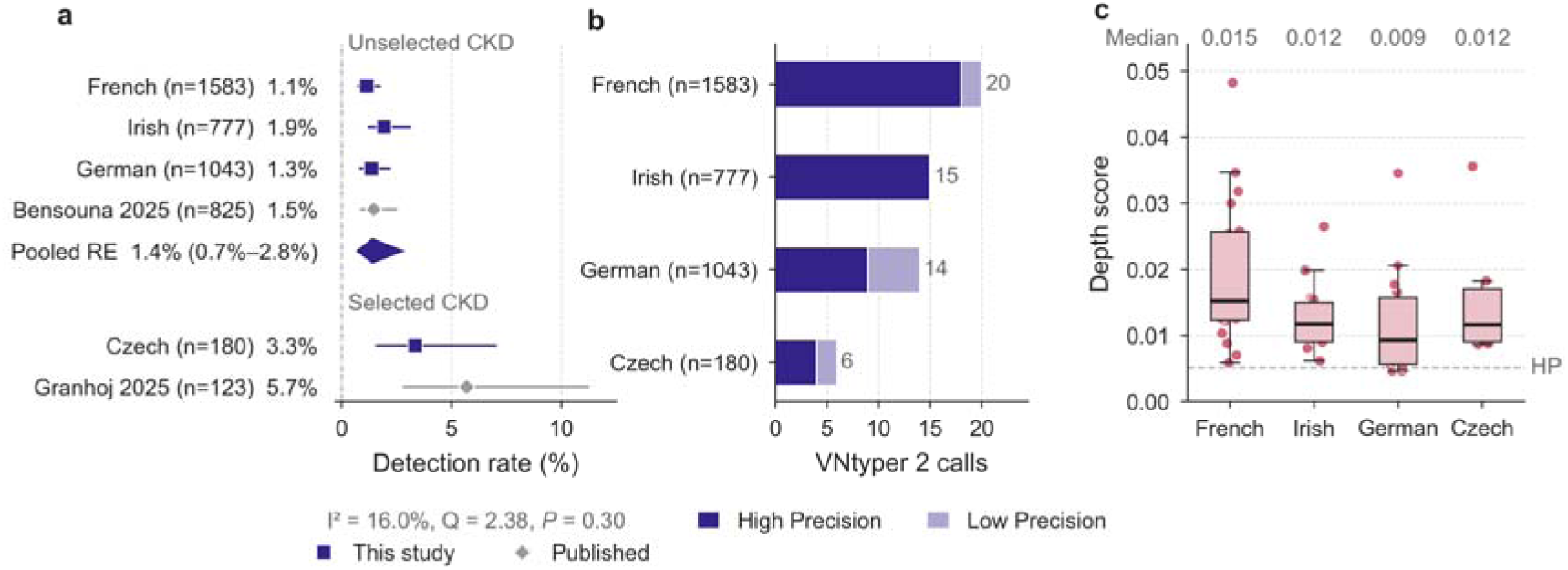
Screening results and prevalence across CKD cohorts. (**a**) Forest plot of *MUC1* VNTR variant detection rates. Cohorts are stratified into unselected CKD programmes from this study (French, Irish, and German), a published unselected French comparator (Bensouna et al.),^8^ and selected CKD cohorts (Czech with prior ADTKD suspicion, and Granhoj et al.^22^ with kidney failure of undetermined aetiology at age 50 or younger). Bensouna et al. are shown as a 59dupC-focused SharkVNTyper comparator; Granhoj et al. detected both 59dupC and atypical non-dupC variants (SNaPshot + VNtyper). Indigo squares indicate cohorts from this study; grey diamonds indicate published comparators. The pooled detection rate for this-study unselected cohorts (diamond) was 1.4% (95% CI 0.7%–2.8%) using a descriptive logit random-effects model with three cohorts. Error bars represent 95% Wilson score confidence intervals. (**b**) VNtyper 2 call breakdown by confidence tier per cohort. Bars show high-precision (dark indigo) and low-precision (light indigo) calls; French low-precision calls are candidate calls and are not included in final positive prevalence estimates. Labels at the bar ends give total VNtyper 2 calls. (**c**) Depth score distributions for final positive calls across cohorts. Box plots show median (dar line) and interquartile range; the dashed line marks the high-precision threshold; median depth scores are listed above the panel. Y-axis censored at 95th percentile; outlier counts annotated. Higher depth score indicate greater confidence in the variant call.

Of the 53 final positives, 44 carried 59dupC (a single-base insertion at position 67 in VNtyper output) and 9 carried atypical non-dupC frameshifts, which occurred in all four cohorts: 3 Czech, 2 German, 3 French and 1 Irish (Supplementary Table S4).

### Towards a testing workflow for ADTKD-*MUC1*

These results support VNtyper 2 as a screening and triage step for *MUC1* VNTR variants within locally validated laboratory workflows when VNTR coverage and sample quality are adequate. VNtyper 2 provides analytical detection, not a clinical diagnosis: positive calls require orthogonal confirmation, and a negative result does not exclude ADTKD-*MUC1*. Because it runs on standard exome alignments with adequate VNTR coverage (Figure 4a), or on raw FASTQ files through SHARK filtering, it fits existing pipelines at no additional assay cost; Twist Exome v2 data aligned with the Burrows-Wheeler Aligner (BWA-MEM) are the best-tested configuration. In 600 downsampling runs of 60 confirmed positive exomes, sensitivity rose from 46% below 50× to 96% at 100× or more GRCh38 VNTR window coverage. The pooled flank depth for 90% sensitivity was 112× (95% CI 71× to 172×), but it depended on the sample: variants called High Precision at full depth reached this sensitivity at 66×, whereas Low Precision variants needed near-full depth (Supplementary Methods S9; Table S11). Laboratories using other capture designs or pipelines should check VNTR coverage and benchmark against orthogonally confirmed samples before treating negative results as informative. We propose an expert-informed two-pathway testing workflow (Figure 5), developed through ADTKD-Net consortium consensus (EJP RD JTC 2023).

**Figure 4:**
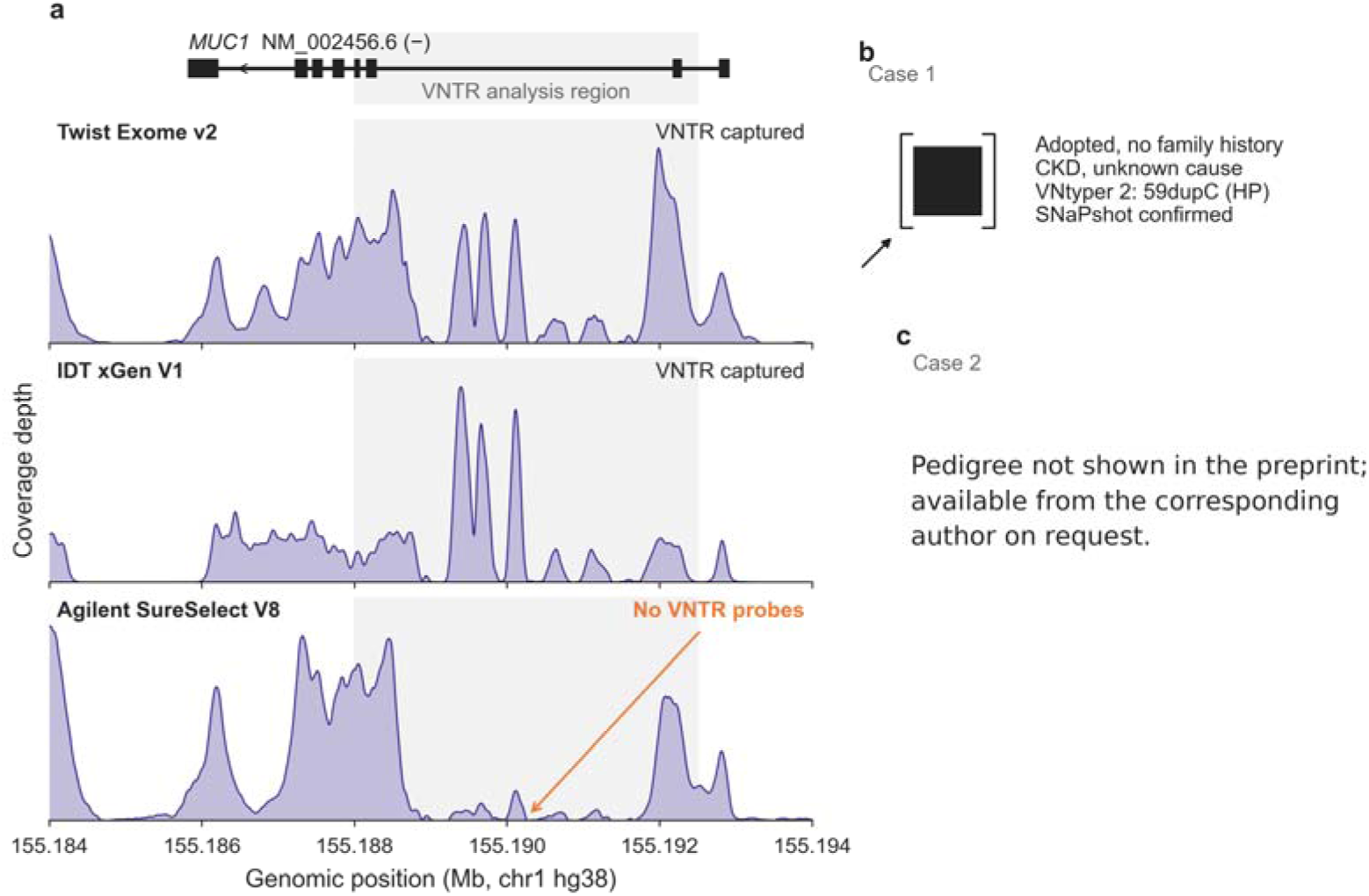
Clinical utility: capture kit coverage and exemplary cases. (**a**) Coverage depth across the *MUC1* locus (chr1:155,184,000-155,194,000, hg38) for three commonly used clinical exome capture designs. The gene track shows UCSC hg38 ncbiRefSeq transcript NM_002456.6. The VNTR region (grey shading) spans approximately 4.5 kb. Twist Exome v2 and IDT xGen V1 [+] provide adequate VNTR coverage through flanking probes and spillover reads. Agilent SureSelect V8 [-], a research exome without VNTR-targeting probes, shows a coverage gap within the repeat region. Profiles are from representative single samples (50-bp rolling mean); absolute depth varies with sequencing depth. Coverage peaks within or near the VNTR reflect capture, alignment, and reference-repeat representation rather than motif-resolved VNTR structure. (**b**) Case 1: an adopted individual with progressive CKD of unknown etiology and no accessible family history. Standard exome analysis identified no pathogenic variants. Retrospective VNtyper 2 analysis detected a canonical 59dupC variant (High Precision), confirmed by SNaPshot. (**c**) Case 2: a family with autosomal dominant CKD; the pedigree is available from the corresponding author on request. External SNaPshot minisequencing had reported the index patient as *MUC1*-negative. VNtyper 2 analysis of the exome data detected a canonical 59dupC insertion, which was subsequently confirmed by the external laboratory on both the original and an independent sample. Without VNtyper 2, the kidney disease in this family would have remained unexplained.

**Figure 5:**
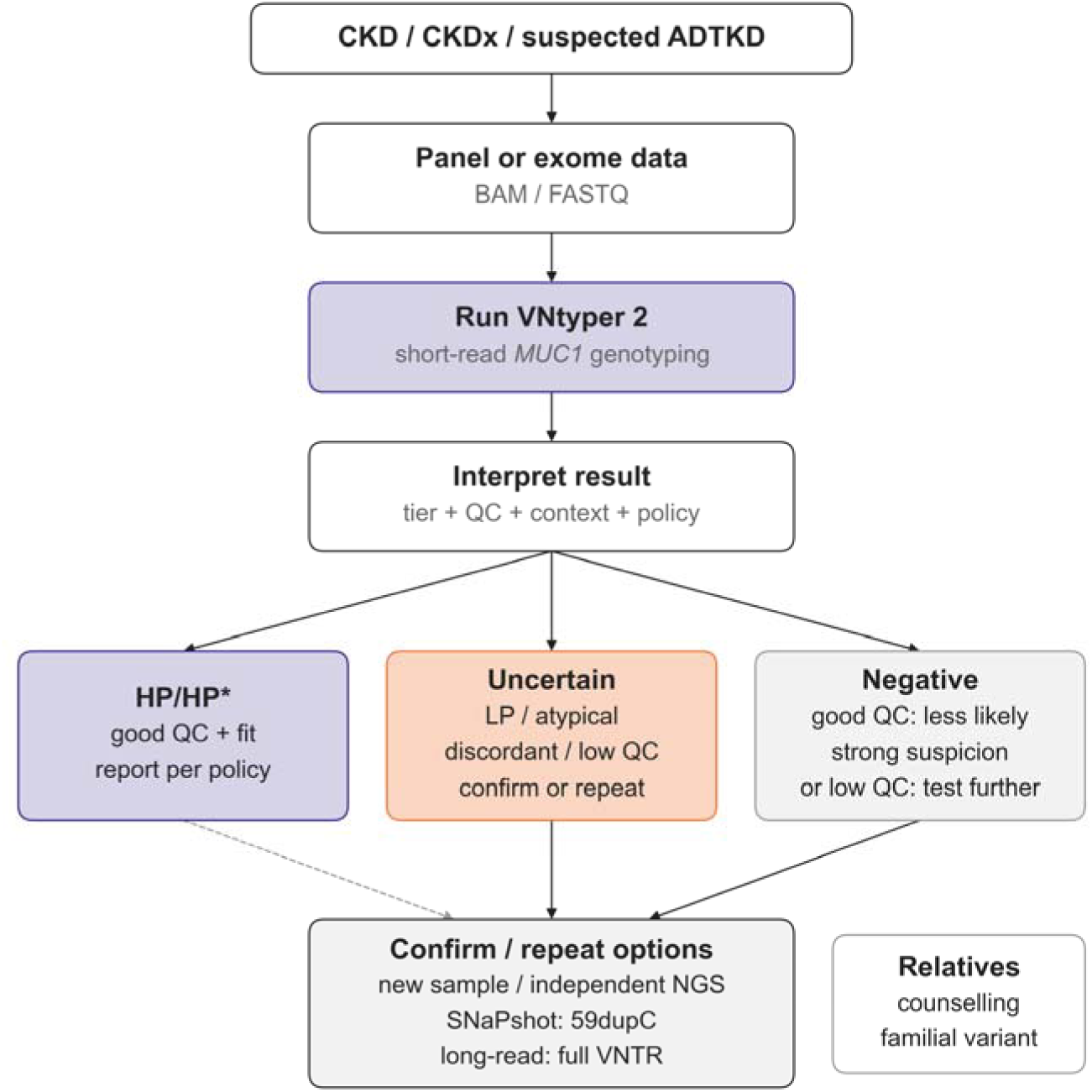
Proposed expert-informed testing workflow for ADTKD-*MUC1*. Patients with CKD, CKD of unexplained cause (CKDx), or suspected ADTKD enter the workflow through standard panel or exome sequencing; VNtyper 2 is run on these data as a computational screening step. Results are interpreted in three tiers: High Precision (HP/HP*), Uncertain (low precision, atypical, discordant, or low QC), and Negative. Confirmation and onward testing depend on confidence tier, variant type, sample quality, clinical context, local validation, and local reporting policy. Confirmation options include 59dupC-targeted probe-extension testing (SNaPshot) and long-read sequencing of the full VNTR. Solid arrows mar expected pathways; the dashed connector from HP/HP* indicates that confirmation is optional and context-dependent rather than mandatory. Predictive testing of relatives should follow counselling and known familial variant testing. HP, high precision; LP, low precision.

In the first pathway, VNtyper 2 runs within pipelines that process panel or exome data for patients with CKD of unexplained cause (CKDx),^26^ the use case our screening supports. Interpretation combines confidence tier, variant type, coverage verdict and clinical concordance. Unflagged High Precision calls in clinically concordant cases are strong candidates, which we recommend confirming orthogonally before clinical reporting unless the laboratory has validated this call type locally. Low Precision calls, atypical or unexpected variants, discordance between Kestrel and code-adVNTR, and suboptimal data should prompt orthogonal confirmation or repeat testing. VNtyper-Online offers the same analysis for research use and workflow evaluation; clinical reporting should use a locally validated implementation, which can be a private deployment.

In the second pathway, patients with strong clinical suspicion of ADTKD-*MUC1* (positive family history, progressive CKD with bland urine sediment, unspecific kidney imaging, and/or hyperuricaemia or gout) also undergo panel or exome sequencing with VNtyper 2 as the initial screen. A negative or low-confidence result does not end the evaluation: SNaPshot minisequencing^12^ rapidly tests for 59dupC, whereas long-read sequencing resolves the complete VNTR, detects non-dupC frameshifts^10^ and localises the variant exactly, which may matter for genotype-phenotype studies. Two cases illustrate this approach: an adopted individual without family history diagnosed from existing exome data (Figure 4b), and a family in whom VNtyper 2 detected a 59dupC insertion that external SNaPshot testing had missed and later confirmed on re-testing (Figure 4c).

## Discussion

We present VNtyper 2 and VNtyper-Online as freely available high-speed tools for *MUC1* VNTR genotyping from short-read sequencing data. Across simulation, orthogonally confirmed clinical validation and retrospective screening of unselected and selected CKD cohorts, VNtyper 2 detected both canonical 59dupC and atypical non-dupC frameshift variants. The pooled detection rate in unselected CKD cohorts was consistent with independent screening data, while selected cohorts showed higher yields, as expected from clinical enrichment.

Most importantly, 9 of 53 final positives (17%) carried atypical non-dupC frameshifts at various positions within the VNTR. PacBio confirmed atypical variants in the Czech validation data, whereas atypical positives in the screening cohorts remain screening findings until confirmed by a VNTR-resolving method. Most atypical variants are invisible to 59dupC-targeted probe-extension assays such as SNaPshot minisequencing and mass spectrometry-based formats,^10–12^ so screening with these assays alone would miss a substantial proportion of affected individuals. Long-read sequencing, whether PacBio SMRT amplicon sequencing^10,13^ or Oxford Nanopore,^14^ resolves the complete VNTR and can confirm any frameshift type, but requires additional DNA, dedicated library preparation, new sequencing runs and specific clinical suspicion, which limits its use for unbiased screening of existing short-read cohorts. Simplified PCR-based assays^27^ and immunohistochemical detection of MUC1-fs^16^ serve specific clinical scenarios, but none matches the scalability and variant breadth of a computational tool applied to existing short-read data. VNtyper 2 fills this gap as a screening and triage step within locally validated workflows.

The clinical value of VNtyper is practical. ADTKD-*MUC1* often resembles more common kidney diseases, and a genetic diagnosis can end years of uncertainty, guide cascade testing and avoid unnecessary invasive procedures. Earlier diagnosis also matters for emerging therapies and cohort building.^2^ Although no disease-specific treatment is approved yet, natural history studies and trial readiness need genotyped patient cohorts, including through registries such as ADTKD-Net (https://www.erknet.org/subregistries/adtkd-registry).

Several aspects of our analysis deserve comment. First, the validation cohorts differed in referral context, variant spectrum, capture design and assembly. This limits direct comparison between them but tests VNtyper 2 in distinct diagnostic settings. Second, no single method is an unambiguous gold standard for *MUC1* VNTR genotyping: PacBio long-read sequencing reconstructs the repeat most completely but may miss variants in extremely long or complex alleles,^10^ and SNaPshot detects only 59dupC. Discordant cases may therefore reflect limitations of either method, which higher-depth SMRT sequencing or MUC1-fs immunohistochemistry^16^ could resolve.

Third, sensitivity falls as the mutated allele lengthens, because one mutated repeat unit makes up a smaller share of a longer array (Supplementary Methods S3). VNtyper 2 therefore estimates the total VNTR length of both alleles and notes long arrays in the report and on VNtyper-Online (Supplementary Methods S12), and running code-adVNTR beside Kestrel, the VNtyper-Online default, recovers most atypical variants that Kestrel misses.

Fourth, sensitivity depended on the sample as much as on depth, so our downsampling series supports no universal minimum coverage (Supplementary Methods S9). Typical clinical genomes fall short: in an internal test, VNtyper 2 detected 1 of 3 confirmed carriers sequenced as genomes at about 40× mean depth, so genome data need local validation before use. The coverage verdict and confidence grade show when a negative call is informative, but adequate coverage alone does not exclude a variant; for clinically suspected ADTKD-*MUC1* with a negative result, we recommend long-read sequencing or, for 59dupC, SNaPshot minisequencing. Laboratories should validate VNtyper 2 on their own capture kit with orthogonally confirmed samples, supported by its calibration module (Supplementary Methods S8). Independent validation in France, Denmark and Japan^8,20,22^ supports the generalisability of our findings, and the ADTKD-Net consortium and opt-in data donation on VNtyper-Online (Supplementary Methods S2) will extend calibration across kits.

In conclusion, VNtyper 2 and VNtyper-Online make *MUC1* VNTR genotyping accessible from existing short-read data at no additional assay cost. By lowering the technical and financial barriers to ADTKD-*MUC1* testing, these tools can help end the diagnostic odyssey for affected families and prepare genotyped patient cohorts for the targeted therapies of tomorrow.

## Disclosure

VJ holds ownership interests in a consulting company providing services in genomics and bioinformatics. This entity had no role in the design, funding, execution, or interpretation of the present study. BP and JoH are employed by Labor Berlin-Charité Vivantes GmbH, a diagnostic laboratory offering genetic testing services for inherited kidney disease, including ADTKD diagnostics. All other authors declared no competing interests.

## Acknowledgments

We thank the patients and families who participated in this study. We gratefully acknowledge the ADTKD-Selbsthilfe patient organisation (https://www.adtkd.de/) for their continued advocacy, support, and engagement with the research community, which has been instrumental in raising awareness of ADTKD and motivating the development of accessible diagnostic tools. We also thank the members of the ADTKD-Net consortium for their contributions to the discussions and consensus that informed the proposed testing workflow; individual non-author contributors are listed in the Supplementary Acknowledgements.

## Data Sharing Statement

VNtyper 2 (https://github.com/vntyper-org/VNtyper; BSD 3-Clause License), VNtyper-Online (https://vntyper.org; source code for the frontend, https://github.com/vntyper-org/vntyper-online-frontend, and backend, https://github.com/vntyper-org/vntyper-online-backend; MIT License) and MucOneUp (https://github.com/vntyper-org/MucOneUp; MIT License) are open source. De-identified per-sample results are available on Zenodo (Supplementary Data S1 to S4; https://doi.org/10.5281/zenodo.20014840). Individual-level sequencing data cannot be shared because of consent and data-protection restrictions; requests for collaborative re-analysis can be addressed to the corresponding author.

## Author Contributions

BP and HS conceived and designed the study, developed the software, performed VNtyper data analysis, and wrote the manuscript. OT and VJ performed VNtyper data analysis. JoH contributed to early VNtyper development and partial data analysis. MŽ, SK, VJ, AP, AV, and HH coordinated and performed *MUC1* VNTR analysis using the long-read SMRT PacBio assay. VJ, AP, and AV analyzed the PacBio sequencing data. CR, EEH, MC, GC, and PC provided Irish samples and clinical data. AJB provided samples, clinical data, and contributed to study design. MW provided samples and clinical data. KUE contributed to study design and acquired funding. SK provided samples, contributed to study design, and acquired funding. CA provided samples, contributed to study design, acquired funding and edited the manuscript. GD, MŽ, and JaH contributed to study design, provided samples, acquired funding, and contributed to writing the manuscript. All authors read and approved the final manuscript.

## Supplementary Material

Supplementary material is available online with this preprint.

## Declaration of Generative AI and AI-assisted technologies in the manuscript preparation process

During the preparation of this work the authors used Anthropic Claude (Opus 4 to Opus 5.5, Sonnet 4), GitHub Copilot, and QuillBot in order to assist with software development, data analysis scripting, figure generation, and to support language refinement and clarity of expression in the manuscript. After using these tools, the authors reviewed and edited the content as needed and take full responsibility for the content of the published article.

## References

1. 1. Bleyer AJ, Živná M, Kidd K, Kmoch S. Autosomal Dominant Tubulointerstitial Kidney Disease – MUC1. In: Adam MP, Feldman J, Mirzaa GM, et al., eds. GeneReviews®. University of Washington, Seattle; 2013. Accessed May 7, 2025. http://www.ncbi.nlm.nih.gov/books/NBK153723/

2. Dvela-Levitt M, Shaw JL, Greka A. A Rare Kidney Disease To Cure Them All? Towards Mechanism-Based Therapies for Proteinopathies. Trends in Molecular Medicine. 2021;27(4):394–409. doi:10.1016/j.molmed.2020.11.008

3. Olinger E, Hofmann P, Kidd K, et al. Clinical and genetic spectra of autosomal dominant tubulointerstitial kidney disease due to mutations in UMOD and MUC1. Kidney International. 2020;98(3):717–731. doi:10.1016/j.kint.2020.04.038

4. Mabillard H, Sayer JA, Olinger E. Clinical and genetic spectra of autosomal dominant tubulointerstitial kidney disease. Nephrology, Dialysis, Transplantation: Official Publication of the European Dialysis and Transplant Association - European Renal Association. 2023;38(2):271–282. doi:10.1093/ndt/gfab268

5. Yamamoto S, Kaimori JY, Yoshimura T, et al. Analysis of an ADTKD family with a novel frameshift mutation in MUC1 reveals characteristic features of mutant MUC1 protein. Nephrology, Dialysis, Transplantation: Official Publication of the European Dialysis and Transplant Association - European Renal Association. 2017;32(12):2010–2017. doi:10.1093/ndt/gfx083

6. Eckardt KU, Alper SL, Antignac C, et al. Autosomal dominant tubulointerstitial kidney disease: Diagnosis, classification, and management—A KDIGO consensus report. Kidney International. 2015;88(4):676–683. doi:10.1038/ki.2015.28

7. Kirby A, Gnirke A, Jaffe DB, et al. Mutations causing medullary cystic kidney disease type 1 lie in a large VNTR in MUC1 missed by massively parallel sequencing. Nature Genetics. 2013;45(3):299–303. doi:10.1038/ng.2543

8. Bensouna I, Robert T, Vanhoye X, et al. Systematic Screening of Autosomal Dominant Tubulointerstitial Kidney Disease–MUC1 27dupC Pathogenic Variant through Exome Sequencing. Journal of the American Society of Nephrology. 2025;36(2):256–263. doi:10.1681/ASN.0000000503

9. Popp B, Ekici AB, Knaup KX, et al. Prevalence of hereditary tubulointerstitial kidney diseases in the German Chronic Kidney Disease study. European journal of human genetics: EJHG. 2022;30(12):1413–1422. doi:10.1038/s41431-022-01177-9

10. Vrbacká A, Přistoupilová A, Kidd KO, et al. Single-Molecule Real-Time Sequencing for *MUC1* Variable Number of Tandem Repeat Variation to Improve Autosomal Dominant Tubulointerstitial Kidney Disease Diagnosis. Journal of the American Society of Nephrology. 2026;37(10):2248–2261. doi:10.1681/ASN.0000001103

11. Blumenstiel B, DeFelice M, Birsoy O, et al. Development and Validation of a Mass Spectrometry–Based Assay for the Molecular Diagnosis of Mucin-1 Kidney Disease. The Journal of Molecular Diagnostics. 2016;18(4):566–571. doi:10.1016/j.jmoldx.2016.03.003

12. Ekici AB, Hackenbeck T, Morinière V, et al. Renal fibrosis is the common feature of autosomal dominant tubulointerstitial kidney diseases caused by mutations in mucin 1 or uromodulin. Kidney International. 2014;86(3):589–599. doi:10.1038/ki.2014.72

13. Wenzel A, Altmueller J, Ekici AB, et al. Single molecule real time sequencing in ADTKD-MUC1 allows complete assembly of the VNTR and exact positioning of causative mutations. Scientific Reports. 2018;8(1):4170. doi:10.1038/s41598-018-22428-0

14. Madritsch S, Horner D, Löwenstern T, et al. Analysis of clinically relevant large tandem repeats using nanopore sequencing. Scientific Reports. 2026;16(1):762. doi:10.1038/s41598-025-30441-3

15. Okada E, Morisada N, Horinouchi T, et al. Detecting MUC1 Variants in Patients Clinicopathologically Diagnosed With Having Autosomal Dominant Tubulointerstitial Kidney Disease. Kidney International Reports. 2022;7(4):857–866. doi:10.1016/j.ekir.2021.12.037

16. Živná M, Kidd K, Přistoupilová A, et al. Noninvasive Immunohistochemical Diagnosis and Novel MUC1 Mutations Causing Autosomal Dominant Tubulointerstitial Kidney Disease. Journal of the American Society of Nephrology: JASN. 2018;29(9):2418–2431. doi:10.1681/ASN.2018020180

17. Živná M, Kidd KO, Barešová V, et al. Autosomal dominant tubulointerstitial kidney disease: A review. American Journal of Medical Genetics Part C, Seminars in Medical Genetics. 2022;190(3):309–324. doi:10.1002/ajmg.c.32008

18. Saei H, Morinière V, Heidet L, et al. VNtyper enables accurate alignment-free genotyping of MUC1 coding VNTR using short-read sequencing data in autosomal dominant tubulointerstitial kidney disease. iScience. 2023;26(7):107171. doi:10.1016/j.isci.2023.107171

19. Park J, Bakhtiari M, Popp B, et al. Detecting tandem repeat variants in coding regions using code-adVNTR. iScience. 2022;25(8):104785. doi:10.1016/j.isci.2022.104785

20. Nagano C, Morisada N, Inoki Y, et al. Clinical use of the VNtyper-Kestrel pipeline for MUC1 variant detection in autosomal-dominant tubulointerstitial kidney disease. Clinical and Experimental Nephrology. 2025;29(9):1192–1199. doi:10.1007/s10157-025-02675-y

21. Kachmar J, Saei H, Morinière V, et al. Phenotypic Heterogeneity of ADTKD-MUC1 Diagnosed Using VNtyper, a Novel Genetic Technique. American Journal of Kidney Diseases: The Official Journal of the National Kidney Foundation. 2025;85(5):603–609.e1. doi:10.1053/j.ajkd.2024.11.010

22. Granhøj J, Lildballe DL, Pedersen KV, et al. MUC1-associated autosomal dominant tubulointerstitial kidney disease: Prevalence in kidney failure of undetermined aetiology and clinical insights from Danish families. Clinical Kidney Journal. 2025;18(1):sfae355. doi:10.1093/ckj/sfae355

23. Audano PA, Ravishankar S, Vannberg FO. Mapping-free variant calling using haplotype reconstruction from k-mer frequencies. Berger B, ed. Bioinformatics. 2018;34(10):1659–1665. doi:10.1093/bioinformatics/btx753

24. Denti L, Pirola Y, Previtali M, et al. Shark: Fishing relevant reads in an RNA-Seq sample. Ponty Y, ed. Bioinformatics. 2021;37(4):464–472. doi:10.1093/bioinformatics/btaa779

25. Popp B, Saei H. MucOneUp: A Simulation Framework for MUC1-VNTR Variant Benchmarking. Published online May 2026. doi:10.64898/2026.05.08.723876

26. Halbritter J, Figueres L, Van Eerde AM, et al. Chronic Kidney Disease of unexplained cause (CKDx): A consensus statement by the Genes & Kidney Working Group of the ERA. Nephrology, Dialysis, Transplantation: Official Publication of the European Dialysis and Transplant Association - European Renal Association. 2025;40(12):2390–2400. doi:10.1093/ndt/gfaf092

27. Fages V, Bourre F, Larrue R, et al. Description of a New Simple and Cost-Effective Molecular Testing That Could Simplify MUC1 Variant Detection. Kidney International Reports. 2024;9(5):1451–1457. doi:10.1016/j.ekir.2024.01.058

