## Supplementary Methods, Figures and Tables for "VNtyper 2 enables open-access short-read genotyping of MUC1 VNTR variants in ADTKD at high-speed"

**Supplementary Data Files 46**

**ADTKD-Net Consortium Acknowledgements 47**

**References 48**

### 1 Supplementary Methods

#### 1.1 S1. VNtyper 2 Pipeline Architecture

##### 1.1.1 Algorithmic Overview

VNtyper 2 detects frameshift variants in the coding VNTR of MUC1 with Kestrel, a k-mer-based caller that compares VNTR reads with the expected repeat motifs without alignment to a linear reference and so avoids mapping bias in this GC-rich repeat.[S1,S2] The pipeline runs in nine steps:

1. **Input validation.** One BAM, CRAM or FASTQ file per run is checked for integrity and format.
2. **Preprocessing.** fastp quality-trims FASTQ input. From BAM or CRAM input, samtools extracts the MUC1 region (chr1:155,158,000-155,163,000 on GRCh37; chr1:155,184,000-155,194,000 on GRCh38) and the unmapped reads, and computes coverage over the VNTR window (chr1:155,160,500-155,162,000 on GRCh37; chr1:155,188,000-155,192,500 on GRCh38). Coordinates are 1-based and inclusive; build and contig naming (UCSC, Ensembl or NCBI RefSeq) come from the file header.
3. **Kestrel genotyping.** Kestrel analyses the reads against the MUC1 VNTR motif reference (k-mer size 20; at most 40 alignment and haplotype states) and writes candidate variants to VCF.
4. **Variant scoring.** A candidate is a frameshift when its alternate-minus-reference length (divided by three, the frame score) is not a whole number. The haplotype count records how many Kestrel haplotype calls support the variant.
5. **Confidence assignment.** Each candidate receives a tier from its depth score and alternate depth (Supplementary Methods S6), with thresholds from the original VNtyper analyses of Twist custom-capture data,[S2] not tuned on the cohorts analysed here.
6. **Flagging.** Configurable rules flag a known 4-bp CGGCA insertion artifact, which is never a call, and variants in conserved motifs with a depth score below 0.4, which are deprioritised.
7. **Variant selection and naming.** A deterministic priority selects one call per sample (Supplementary Methods S6). The call is named on the canonical X repeat unit in the VNTR nomenclature of the literature (for example 59dupC), placed by the HGVS rule for equivalent positions, with a naming tier and the window of equivalent positions.
8. **Cross-matching.** When code-adVNTR runs, its calls are matched to Kestrel's by allele change and variant type.
9. **Report.** An HTML report shows the calls with an interactive IGV.js read view; cohort aggregation is supported.

##### 1.1.2 code-adVNTR

VNtyper 2 optionally runs code-adVNTR,[S3] a profile hidden Markov model genotyper, from a maintained fork (https://github.com/vntyper-org/adVNTR) that reduces false positives, compiles with GCC 14, decodes reads in parallel and runs in its own environment. VNtyper 2.0.47 pins adVNTR v2.4.1. The step took a median of 20.0 to 28.4 s per GRCh38 exome (Supplementary Methods S7), and VNtyper-Online runs it by default.

##### 1.1.3 VNTR Coverage Reporting

Because low coverage limits what a negative result says, VNtyper 2 grades each sample PASS, REDUCED or FAIL from mean VNTR coverage and uncovered fraction (limits in Supplementary Methods S9). The limits differ by build because the GRCh37 reference holds fewer repeat units, which raises the mean VNTR-window depth of the same sample. VNtyper 2 also reports depth over the repeat array itself (chr1:155,161,000-155,161,810 on GRCh37; chr1:155,188,530-155,192,010 on GRCh38) and the flank depth, the mean depth over 190 bp of unique sequence on each side, which is comparable between builds.

##### 1.1.4 Sample Confidence Grade

Each report prints one of six sample-level confidence grades, assigned by an explicit rule table from the Kestrel result, flags, agreement between Kestrel and code-adVNTR, and the coverage verdict: “Finding, corroborated” (Kestrel call confirmed by code-adVNTR); “Finding”; “Finding, with caveats” (flagged call, or coverage not measured or mostly uncovered); “No finding”; “No finding, inconclusive” (FAIL or unmeasured coverage); and “No result” (genotyping incomplete). Next to the grade, a negative Kestrel result with a frame-valid candidate below the reporting threshold reads “Negative, candidate below threshold”, and a negative at REDUCED coverage carries a note that a weak variant signal can be missed at this depth. Subthreshold candidates also occur in true negatives, so they do not lower the grade.

##### 1.1.5 VNTR Length Estimate and Long-Array Note

Kestrel sensitivity falls as the mutated allele lengthens (Supplementary Methods S3), so VNtyper 2 estimates the total VNTR length of both alleles from the read-depth pattern across the repeat, with a GRCh38 model trained on exomes with PacBio allele lengths. Above a configurable cutoff (150 repeats by default), the report notes that sensitivity may be reduced and a negative result may need orthogonal testing. The research-use estimate is shown with its typical error of about 14 repeats; a normal total does not exclude one long allele. Supplementary Methods S12 describes the model and its validation. VNtyper-Online shows the estimate and the note with each result.

###### 1.1.6 Software Engineering, Interface and Deployment

Continuous integration runs Ruff linting, mypy type checking and unit, integration and container tests on Python 3.9 to 3.12. Thresholds, coordinates and scoring parameters live in JSON configuration files, and every run records per-step timestamps, file checksums and tool versions. The command line covers genotyping, reference installation, reporting, multi-sample aggregation with optional pseudonymisation, calibration (Supplementary Methods S8) and submission to VNtyper-Online. VNtyper 2 installs with pip or conda or runs from a Docker or Apptainer container; a Snakemake workflow processes large cohorts on clusters.

###### 1.1.7 Software Environment

All clinical and simulation analyses used VNtyper 2.0.47 in a pinned container with Kestrel, SHARK 1.2.0, BWA-MEM 0.7.18, samtools 1.20, fastp 0.23.4, OpenJDK 11.0.23, Python 3.12.13, bcftools 1.21 and pysam 0.22.1.

#### 1.2 S2. VNtyper-Online Web Application

##### 1.2.1 Frontend

VNtyper-Online (https://vntyper.org) runs VNtyper 2 without installation. As in other client-side WebAssembly genomics tools,[S4] a WebAssembly build of samtools (BioWasm Aioli) extracts the MUC1 region, typically a few kilobytes, in the browser, and only this region is uploaded. In normal mode, the browser also extracts the unplaced unmapped reads of the BAM file, which local runs add to the Kestrel input. The plain-JavaScript frontend supports hg19, hg38, GRCh37 and GRCh38, processes samples in parallel and includes a tutorial.

##### 1.2.2 Backend and Cohort Mode

A FastAPI service with Celery workers, a Redis broker and per-client rate limiting runs the jobs. It is deployed with Docker Compose behind an nginx reverse proxy, serves all traffic over HTTPS/TLS and received an A+ rating from an independent security assessment; the same setup serves private institutional deployments. Uploaded regions are deleted immediately after analysis and results after seven days. Users can group samples into a passphrase-protected cohort, follow its progress, download all results together and receive an email on completion.

##### 1.2.3 Opt-in result donation

An opt-in step lets users donate one record per sample to a shared calibration set across capture kits. The browser reduces each result to binned and enumerated values (VNtyper version, coverage verdict, flank and repeat-array depth, unflagged or flagged call, confidence tier, depth score with fine bins near the reporting threshold, 59dupC or other variant), to which the user adds capture kit, platform class, orthogonal result and confirmation method. No sequence, file name, variant name or sample identifier leaves the browser. The step stays off on the public instance until the participating universities have completed legal and ethics review; this study reports no donated data.

##### 1.2.4 Online versus local execution

Identical MUC1 region slices of 30 German CeRKiD exomes (14 positive) and 59 reference-standard exomes, cut with the browser's samtools command, were run on the public VNtyper-Online instance and with the same VNtyper 2.0.47 container locally. Calls, confidence tiers, flags, depth scores and coverage verdicts were identical in 89 of 89 samples. Median server computation was 6.3 s per sample (local 4.3 s), and the single public worker processed 340 to 350 jobs per hour.

#### 1.3 S3. MucOneUp Simulation Framework

MucOneUp 0.53.0 (https://github.com/vntyper-org/MucOneUp),[S5] run with its default configuration, generated simulated MUC1-VNTR sequencing data with known ground truth.

##### 1.3.1 Preregistered held-out design

The design was registered before any genotyping run and used seeds disjoint from all development runs; the held-out set was never used to tune VNtyper 2, MucOneUp or the analysis code.

**Natural-length simulation (primary benchmark).** 500 matched wild-type and mutated pairs for 59dupC and 500 pairs for ten atypical non-dupC frameshifts, 50 per type: insG, dupA, delinsAT, insCCCC, insC_pos23, insG_pos58, insG_pos54, insA_pos54, delGCCCA and ins25bp.[S2,S6,S7] Each allele length was drawn from a normal distribution (mean 50, SD 17 repeats) truncated to 35–105 repeats by resampling, within the range of PacBio-resolved allele lengths,[S7] and passed to MucOneUp as a fixed length. All 2000 BAMs were also downsampled with samtools to 75%, 50%, 25%, 12.5% and 6.25%.

**Long-allele simulation.** 100 59dupC and 100 atypical pairs (10 per type), with the variant on an allele drawn uniformly from 105–140 repeats and the other allele drawn as in the natural-length simulation. The long-allele simulation is never pooled with the natural-length simulation.

##### 1.3.2 Sample generation

For each pair, MucOneUp built two VNTR haplotypes and introduced the variant into one haplotype of the mutated sample only. Illumina reads were simulated at 150× with the shipped read model and exome-like downsampling (Twist Exome v2 VNTR-to-flank coverage ratio), aligned with BWA-MEM to GRCh38 and sorted. insG_pos54 and insA_pos54 arise on repeat units (bJ, aH) that random MucOneUp chains never contain, so a unit one mismatch away (D or E for bJ, B for aH) was chosen from the seed and converted. Two pairs lacking such a unit used the next-closest units (preregistered amendment based only on haplotype structure). All 1000 N pairs and 200 L pairs were analysed (Data S1).

##### 1.3.3 VNtyper 2 execution and classification

We ran VNtyper 2.0.47 on GRCh38 in three arms: Kestrel in normal mode on all BAMs and titration fractions, Kestrel plus code-adVNTR, and fast mode. A sample was positive only with an unflagged call of a tier other than Negative (true positive if mutated, false positive if wild type), for Kestrel and code-adVNTR alike. Flagged calls are never positive and never false positives, and 4-bp insertion artifacts are filtered.

##### 1.3.4 Performance metrics

Sensitivity and specificity per variant group and type have Wilson 95% confidence intervals.[S8,S9] Length dependence was assessed by mutated-allele length bin and by logistic regression of detection on mutated-allele length. The union of Kestrel and code-adVNTR (positive if either made an unflagged call) and depth score by length are exploratory.

##### 1.3.5 Additional simulation results

Supplementary Figures S1–S3 show detection by allele length, variant type and coverage and the depth analysis of the natural-length simulation, and Supplementary Figure S4 the length effect in the long-allele simulation. Detection fell with mutated-allele length: the odds ratio per 10 additional repeats was 0.48 (95% CI 0.32 to 0.7) for 59dupC and 0.69 (0.58 to 0.82) for atypical variants, and in the long-allele simulation 59dupC sensitivity fell from 74.1% at 105–115 to 46.2% at 126–140 repeats (Supplementary Table S1b). The cause is dilution: a frameshift changes one repeat unit, so in a longer allele the mutant k-mers make up a smaller share of the repeat reads, and in detected 59dupC samples the median depth score fell from 0.0218 at 35–60 to 0.0103 at 86–105 repeats. code-adVNTR and Kestrel complemented each other by variant type (Supplementary Table S1a); either method together reached 93.8% sensitivity for atypical variants (Kestrel alone 82.0%) with no false positive among 1000 wild-type samples. Fast mode matched normal mode; simulated BAMs lack the unmapped reads it skips, so this arm says little about real exomes.

#### 1.4 S4. Cohort Descriptions and Sequencing Details

Supplementary Table S2 summarises all cohorts; Supplementary Methods S3 describes the simulated data.

##### 1.4.1 Gold Standard Cohort (French and German)

The French-German gold standard cohort comprised 59 samples. The 40 French exomes from the Imagine Institute in Paris entered after 59dupC-targeted SNaPshot screening and were assessed independently by PacBio long-read sequencing, which confirmed 32 as positive and 8 as negative; one sample (FR-GS-006) had an uninterpretable SNaPshot result and a negative PacBio call. The 19 German samples are familial cases from published ADTKD-MUC1 families, confirmed 59dupC-positive by SNaPshot minisequencing[S10,S11] and sequenced with Twist Exome v2 at Labor Berlin.

##### 1.4.2 Validation Cohort Design and Heterogeneity

The validation cohorts were assembled from available orthogonally characterised material, not as a balanced population sample, and VNtyper 2 was not used to select samples for reference testing. The French-German cohort was enriched for SNaPshot-confirmed 59dupC cases, and all 51 of its reference positives carried canonical 59dupC. All 15 atypical non-dupC reference positives came from the Czech-US cohort of unresolved high-suspicion referrals sent for long-read resolution (variant classes in Supplementary Table S3). Sensitivity and specificity are therefore validation-cohort estimates, and the Fisher's exact comparison of cohort sensitivities describes heterogeneity without attributing it to one factor. The French-German cohort detected 50 and missed 1 reference positives and the Czech-US cohort detected 27 and missed 7 (Fisher's exact $P = 0.006$); for ground-truth 59dupC alone, the French-German cohort detected 50 of 51 and the Czech-US cohort 16 of 19 ($P = 0.058$). Within the Czech-US cohort, detection did not differ clearly between 59dupC and atypical non-dupC variants (16/19 versus 11/15; $P = 0.67$). Supplementary Figure S5 shows the Czech-US depth scores and Supplementary Figure S6 the per-cohort confusion matrices.

Capture was uniform within each cohort. The French-German set used Twist Exome v2: 34 French exomes sequenced at Labor Berlin and the 19 German exomes were aligned with BWA-MEM to GRCh38, and six French exomes sequenced at Imagine with DRAGEN/DRAGMAP to GRCh37. The Czech-US cohort used KAPA HyperExome V2 (Roche) exomes aligned with BWA-MEM to GRCh37. Sequencing platforms (Supplementary Table S2) were Illumina NovaSeq X for the 34 French exomes from Labor Berlin, NextSeq 500/550 for the six French GRCh37 exomes, NovaSeq 6000 for the German exomes and NovaSeq 6000 or NovaSeq X for CeRKiD, all with 2 × 151 bp reads; the Czech-US exomes were sequenced on the NovaSeq X Plus with 2 × 159 bp reads, and the Renome panels with 2 × 150 bp reads on the NextSeq 500 or

MiSeq.[S2] The platform of the Czech screening and Irish exomes was not available.

##### 1.4.3 Screening Cohorts

We analysed four screening cohorts from CKD referral programmes: French targeted panels (1583 samples from the Renome study at the Imagine Institute, restricted to samples sequenced after the publication cutoff of Kachmar et al. 2025 to avoid overlap with reported data),[S12] Czech exomes (180 samples), Irish exomes (777 samples from the Irish Kidney Gene Project, captured with Twist Comprehensive Exome with the Twist Mitochondrial Panel for 727 and CeGaT ExomeXtra v6 for 50; sequencers and read lengths changed over the collection years), and German exomes (1043 samples from the Charité Center for Rare Kidney Diseases, CeRKiD; calls in Data S2). Supplementary Table S4 stratifies the final calls by variant and cohort, and Supplementary Figure S7 shows their depth scores.

##### 1.4.4 Ethics

All cohorts were collected under local institutional review board approval with written informed consent, in accordance with the Declaration of Helsinki.[S13] The Charité Ethics Committee approved the German gold standard and CeRKiD screening samples (EA4/066/21), and the ethics committee of the Friedrich-Alexander University Erlangen-Nürnberg approved the German/Erlangen cohort material (251_18 B). The Ethics Review Board of Beaumont Hospital approved the Irish screening cohort (REC 19/28). The French and Renome cohorts were collected under the Imagine Institute Biocollection framework (DC-2020-3994; IRB 00011928). The Ethics Committee of the General University Hospital and First Faculty of Medicine, Charles University, approved the Prague and Czech screening cohorts. The family shown in main-text Figure 4c underwent routine genetic diagnostics; written consent permitted secondary research use and publication of de-identified clinical and genetic information.

#### 1.5 S5. Orthogonal Validation Methods

##### 1.5.1 SNaPshot Minisequencing

We used the 59dupC assay of Ekici et al. with the M13-tagged primers of Saei et al..[S2,S10] MwoI digests wild-type 7C repeats before and after amplification of the protected MUC1 VNTR fragments, which enriches repeat units carrying the canonical 8C insertion. M13-tagged amplicons were purified with ExoSAP and AMPure XP beads and analysed by SNaPshot probe extension and capillary electrophoresis. A sample was positive when the 59dupC extension product appeared at the expected fragment length. The M13 tags allowed Sanger sequencing when variants affecting the MwoI site needed clarification. Variants elsewhere in the VNTR need long-read sequencing, which resolves the full array including atypical non-dupC variants.

##### 1.5.2 Long-Read Sequencing

Long-read MUC1 VNTR sequencing followed the SMRT workflow of Vrbacká et al..[S7] Blood genomic DNA was PCR-amplified across the VNTR as described.[S6] Pooled products of about 1.5-6 kb were cleaned with SPRI magnetic beads, quantified by Qubit fluorometry, sized on the Agilent 5200 Fragment Analyzer, converted to SMRTbell Express Template Prep Kit 2.0 libraries and sequenced on the PacBio Sequel I.

SMRT Link v8.0 generated circular consensus reads, which PacMUC1 mapped to 120 synthetic MUC1 VNTR references varying the number of canonical X repeats between fixed pre-repeat and after-repeat blocks. Reads unsuitable for allele reconstruction were removed (manual IGV review when needed), and allele lengths came from samtools idxstats read-depth peaks. Clair/Clair3 variant calls and bcftools built allele consensus sequences, which custom Python code split into 60-bp repeat units with Kirby/Wenzel labels to summarise allele structure.[S7,S11,S14] Allele-structure and frameshift calls required at least 10× read support; frameshift signals below this were classified as inconclusive.

#### 1.6 S6. Benchmarking Details

##### 1.6.1 Performance Metrics

Sensitivity and specificity are the proportions of reference-positive and reference-negative samples classified correctly; PPV, NPV and F1-score follow standard formulas. Proportions have Wilson score 95% confidence intervals.[S8,S9] Ground truth was the MucOneUp label for simulated data and SNaPshot or long-read sequencing for clinical cohorts (Data S3). PPV and NPV depend on prevalence and ascertainment, so we report them only as descriptors of each validation cohort.

For screening prevalence, final positives followed cohort-specific interpretation of the confidence tiers. Two Low Precision (LP) 59dupC-like calls in the French targeted panels had low alternate-read support and no orthogonal confirmation; they stayed candidates outside final counts and pooled estimates. LP calls from exome cohorts were final calls, subject to clinical review or confirmation. The pooled detection rate in unselected screening cohorts used the final positive calls of the French Renome, Irish and German cohorts in a logit-scale random-effects single-proportion meta-analysis with DerSimonian-Laird $\tau^2$ and a Hartung-Knapp-Sidik-Jonkman confidence interval.[S15–S17] Cochran's Q, $I^2$, $\tau^2$ and a 95% prediction interval summarise between-cohort heterogeneity; with three cohorts, these estimates are descriptive (Supplementary Table S5).

##### 1.6.2 Confidence Scoring

Each call receives one of four tiers from empirically derived thresholds.[S2] The depth score is the ratio of estimated alternate allele depth to estimated depth in the active VNTR region.

- **Negative.** Depth score below 0.00469. No variant is reported.
- **Low Precision.** Depth score between 0.00469 and 0.00515, or alternate allele depth $\leq 20$, or active region depth $\leq 200$. Orthogonal confirmation is recommended.
- **High Precision.** Depth score $\geq 0.00515$ and alternate allele depth between 21 and 99.
- **High Precision*.** Depth score $\geq 0.00515$ and alternate allele depth $\geq$ 100, indicating strong support from multiple independent k-mer assemblies.

When several variants pass, the pipeline reports one, prioritising highest tier, unflagged status, highest depth score, highest haplotype count and lowest VNTR position.

High Precision and High Precision* calls in adequately covered samples can support diagnosis in concordant clinical or familial settings under local laboratory policy. Low Precision calls, atypical or unexpected variants, discordant internal calls, and negative results despite strong clinical suspicion should prompt repeat analysis or orthogonal testing.

##### 1.6.3 SHARK Preprocessing

SHARK extracts reads from raw FASTQ files by Bloom filter-based k-mer matching,[S18] as in the SharkVNTyper workflow for direct FASTQ screening of exomes.[S19] For users without aligned BAM files, VNtyper 2 can run SHARK upstream to pass MUC1-matching paired-end reads to alignment and Kestrel. Our analyses used aligned data.

#### 1.7 S7. Comparison with the Original VNtyper

##### 1.7.1 Design

We compared both versions as whole pipelines on the same BAM files of the three reference-standard cohorts: French (40), German (19) and Czech-US (83), 142 in total. VNtyper 1.3.0, the last release of the original tool, ran from its published container with Kestrel only; VNtyper 2.0.47 ran with default settings, including code-adVNTR. For between-version concordance without a reference standard, we also ran the full Czech exome set (263 samples), which contains the Czech-US samples. Kestrel calls were the primary result for both versions, and intervals are Wilson score 95% intervals.

##### 1.7.2 Mapping calls between versions

VNtyper 1.3.0 has no flag column, so every High Precision, High Precision* or Low Precision row counts as a call; for VNtyper 2 only unflagged calls are positive. The 4-bp insertion artifact rule (alternate allele exactly 4 bp longer than the reference, here C>CGGCA and C>CGGCT) applies to both; for VNtyper 1.3.0 we applied it afterwards and also report results without it (Supplementary Table S6). Variants were matched across the two position schemes (59dupC is POS 7 in VNtyper 1.3.0 and POS 67 in VNtyper 2).

##### 1.7.3 Results

With the artifact rule applied to both, the two versions gave identical counts in every cohort (Supplementary Table S6), with pooled sensitivity of 90.6% (95% CI 82.5%–95.2%) and the same 8 missed positives. Sample status agreed in 141 of 142 reference-standard samples, in 259 of 263 full Czech exomes and in 1574 of 1583 Renome targeted panels. In the exomes, every difference was a VNtyper 1.3.0 call that VNtyper 2 filtered as a 4-bp insertion artifact or flagged; in the panels, the 9 calls unique to VNtyper 1.3.0 were Low Precision calls below the VNtyper 2 reporting floor. VNtyper 2 made no positive call that VNtyper 1.3.0 missed. Both versions detect the same variant classes; VNtyper 2 reports one selected call per sample in VNTR nomenclature. Without the artifact rule, VNtyper 1.3.0 specificity falls from 98.2% to 96.5%. Supplementary Table S7 lists the other differences between the versions. FR-GS-006, reference negative, was called by both versions and by code-adVNTR in two independent sequencing runs; it was negative by PacBio and is counted as a false positive (Supplementary Table S8).

##### 1.7.4 Runtime and scalability

We timed both versions on exclusive cluster nodes of one CPU model with 8 threads per run, on 20 CeRKiD MUC1 region slices (half screening positives; 5 for the slow VNtyper 1.3.0 code-adVNTR arm). To let VNtyper 1.3.0 code-adVNTR run on GRCh38 slices, every arm used read pairs with both mates inside the region (median 99.0% of reads retained). Ratios are medians of paired per-sample ratios.

Kestrel-only genotyping took a median of 9.1 s with VNtyper 1.3.0 and 11.8 s with VNtyper 2.0.47. With code-adVNTR, VNtyper 1.3.0 took 56 min per sample on a single core, and VNtyper 2.0.47 30.5 s, 107-fold less wall time and 33-fold less CPU time; four parallel samples on one 16-core node gave 910 samples per hour. In the cohort runs, code-adVNTR added a median of 20.0 to 28.4 s per exome. In the 777 Irish exomes, code-adVNTR called 13 of the 15 Kestrel positives with the same variant, flagged its 4 additional calls for low coverage, and made no unflagged call that Kestrel missed.

#### 1.8 S8. Threshold Robustness and Local Calibration

##### 1.8.1 Score and replay

Before computing any curve, we defined one score per sample: the highest depth score among frame-valid, motif-filtered, unflagged Kestrel candidates that are not 4-bp insertion artifacts, or 0 if none exists. We replayed the VNtyper 2.0.47 decision rule (confidence rule table, call gates and variant selection) on the stored per-candidate Kestrel tables of all 142 reference-standard samples. The replay reproduced every benchmark call at the shipped values (Supplementary Table S6), so a score at or above t is exactly the call VNtyper 2 would make with its reporting floor at t.

##### 1.8.2 Sweep

We swept the depth-score threshold from 0.0020 to 0.0100 in steps of 0.0001 and computed sensitivity and specificity pooled and by cohort and capture kit, by tertile of flank depth, and by coverage QC verdict. ROC and precision-recall areas under the curve have class-stratified bootstrap 95% confidence intervals (2000 resamples, fixed seed). Counting rules were those of Supplementary Methods S7, with FR-GS-006 as a false positive. Sweeping the alternate-depth and active-region depth values one at a time through the full replay showed that, as shipped, they only split confidence tiers and decide no call.

##### 1.8.3 Results

The pooled ROC area under the curve was 0.978 (95% CI 0.953–0.997; strata in Supplementary Figure S8 and Table S9). Pooled sensitivity and specificity were unchanged from 0.0046 to 0.0051 and within 5 percentage points from 0.0036 to 0.0057. Lowering the threshold recovered 3 of 8 false negatives

(Supplementary Table S10) without a false positive; the next 3 cost 4 to 7 false positives, and 2 had no candidate at any threshold. The cost depended on the kit: all 8 French negatives (Twist Exome v2) carried a low-level 59dupC candidate, whereas 48 of 49 Czech-US negatives (KAPA HyperExome V2) had none. Strata are small and kit is confounded with cohort, so these analyses are exploratory, and we selected no new threshold from them.

##### 1.8.4 Local calibration

Because the trade-off differs between kits, VNtyper 2 ships no kit-specific thresholds. Its calibration commands replay the Kestrel and code-adVNTR decision rules on a laboratory's orthogonally confirmed positives and negatives, select cutoffs on training folds and report held-out sensitivity, specificity and ROC operating points; a derived profile applies only when chosen explicitly, and the packaged defaults never change. We recommend a local profile only if it beats the defaults on held-out samples from the same assay.

#### 1.9 S9. Coverage-Dependent Sensitivity

##### 1.9.1 Design

We downsampled confirmed positive GRCh38 exomes in three series (VNtyper 2.0.47). The primary gold series used the full exomes of 27 French and 19 German gold standard positives, each downsampled once with samtools to 9 fractions (10% to 90%) and run in normal mode with code-adVNTR; the full-depth benchmark run completed each sample (460 runs). Full exomes keep every unmapped read that VNtyper adds to the Kestrel input. FR-GS-006, negative by PacBio, was excluded. The Berlin series used the GRCh38 MUC1 region slices of 14 confirmed positive German CeRKiD exomes, with 3 replicate seeds per fraction plus the full slice (392 runs; Kestrel only). As a sensitivity analysis, we downsampled the region slices of the same 46 gold positives in the Berlin design (1288 runs). The primary analysis pooled the gold full-exome and Berlin series (60 samples). Downsampling kept read pairs together, with fractions nested within a seed. Detection required an unflagged positive Kestrel call; no flagged calls or 4-bp insertion artifacts occurred. Depth was recorded as flank depth (Supplementary Methods S1) and as the GRCh38 VNTR window mean used by the coverage verdict.

##### 1.9.2 Analysis

We fitted a logistic generalised estimating equation (GEE) of detection on log2 depth with an independence working correlation, obtained 95% confidence intervals from 2000 cluster bootstrap resamples over samples (fixed seed), and report the depth at which modelled sensitivity reaches 90% and 95%. Because full-depth detection separated two groups of samples, we also report post hoc strata by full-depth confidence tier (a sample missed at full depth belongs to neither). To weight samples equally, the pooled analysis used one run per sample and fraction (the first seed of each slice series), with all seeds as a sensitivity analysis. All samples are positive, so these series estimate sensitivity only; specificity at low depth comes from the simulation titration (Supplementary Methods S3).

##### 1.9.3 Results

Of 600 pooled runs, 482 detected the variant, including every full-depth run except the known German false negative (Supplementary Table S11, Figure S9). Pooled flank depth for 90% sensitivity was 112× (71× to 172×). The pooled curve averages two kinds of sample: the 51 samples called High Precision at full depth reached 90% at 66×, whereas the 8 called Low Precision, all German, were detected in 37 of 80 runs, mostly near full depth. Full exomes and region slices agreed (90% at 97× and 82× over the gold positives), so the extra unmapped reads of full exomes did not shift detection.

These data support coverage-dependent sensitivity, not a universal minimum coverage. The coverage verdict limits (Supplementary Methods S1) are a reporting policy along this gradient, not validated cut-offs; laboratories should confirm them on their own data (Supplementary Methods S8).

#### 1.10 S10. Performance by Cohort, Capture Kit, and Depth

We stratified the 85 reference-positive and all reference-negative benchmark samples by cohort and capture kit, flank depth band, coverage QC verdict, assembly and reference variant class (Supplementary Table S3), with the counting rules of Supplementary Methods S7 and FR-GS-006 as a false positive. Confidence intervals are exact (Clopper-Pearson) and differ slightly from the Wilson intervals of the main text. To separate kit from depth, we fitted Firth-penalised logistic models of detection among reference positives with cohort and kit as one factor (reference level Czech-US, KAPA HyperExome V2), alone (M1) and with log2 flank depth relative to 100× (M2), and depth alone within the Czech-US cohort (M3). Intervals are profile penalised-likelihood 95% CIs; estimates matched the R package logistf. Each cohort used one kit, so this term also carries centre, library preparation, assembly (all Czech-US samples are GRCh37) and variant spectrum (all atypical positives are Czech-US): it is an association, not a kit effect. With 8 false negatives and two cohorts carrying reference negatives, a mixed model is not identifiable, so each model has at most one covariate besides this factor. Sequencing platform coincides with cohort and assembly and is reported descriptively. Per-sample values are in Data S4.

Twist Exome v2 reached 98.0% sensitivity (50/51) and KAPA HyperExome V2 79.4% (27/34). Depth adjustment left the cohort-and-kit odds ratio almost unchanged (16.5, 95% CI 1.8 to 2191), and each doubling of flank depth carried an odds ratio of 2.07 (0.61 to 8.06). Of the 7 KAPA false negatives, 4 carried atypical frameshifts. At adequate depth, 5 of 6 misses carried a candidate just below the reporting threshold, the situation that per-kit local calibration addresses (Supplementary Methods S8).

In the 58 reference-standard positives with a PacBio-sized mutated allele, sensitivity was 92.9% below 60 repeats, 84.2% from 60 to below 80 and 84.0% at 80 or more (90.9%, 76.9% and 70.0% in the Czech-US cohort; Supplementary Table S3), a descriptive trend in the same direction as the simulation (Supplementary Methods S3).

#### 1.11 S11. Paired-Kit Experiment

Four PacBio-confirmed carriers missed on KAPA HyperExome V2 exomes, three from the Czech-US reference standard and one from the Czech screening cohort, were resequenced from the same DNA with Twist Exome v2 at Labor Berlin. Both exomes were analysed with VNtyper 2.0.47 at default settings (Supplementary Table S12). Chromosome 1 SNV genotypes, matched across assemblies by flanking sequence, confirmed every pair (99.5% to 99.8% genotype concordance within a pair, 65% to 70% between individuals). Both exomes of the Czech-US carriers came from the NovaSeq X; the KAPA exome of the screening carrier came from a NovaSeq 6000.

On Twist, VNtyper 2 called 3 of the 4 carriers at high precision; CZUS-009 scored 0.00467, just below the reporting threshold, which the report states as a subthreshold signal. On KAPA, mutant k-mers were present at the Twist rate, yet no carrier received a call of the PacBio variant at adequate coverage. The three Czech-US KAPA exomes carried 22% of VNTR bases below Q20 against 10% in their Twist pairs, and only 13% of their VNTR reads were nearly free of low-quality bases, against 55%. These errors recur in the GC-rich repeat: the KAPA reads carried about 1.7 times more recurrent non-reference k-mers, mostly C>T and T>C substitutions. Kestrel follows a bounded number of haplotype paths per active region, and each recurrent error opens a competing path, so the variant path is dropped or placed on a stretch with a larger active region and lower depth score. It also moves CZUS-009 on Twist across the threshold depending on which reads are sampled. The KAPA exome of the screening carrier had Twist-level base quality but 771 VNTR reads against 2245 on Twist, and its coverage verdict was FAIL.

This mechanism also explains why simulated sensitivity rose slightly when coverage was halved (Supplementary Table S1): at full depth, more recurrent errors pass the k-mer count and compete with the variant path. VNTR base quality is therefore a second input property, besides coverage, that sets how much a negative call says; laboratories whose libraries carry many low-quality bases in GC-rich regions should calibrate locally (Supplementary Methods S8).

#### 1.12 S12. VNTR Length Estimate

##### 1.12.1 Model

We trained the length model on 76 Twist exomes, 42 German and 34 French, whose diploid VNTR length PacBio sequencing had measured (66 to 175 repeats, counting the nine invariant units of each allele). Thirteen features describe the MUC1 locus in the GRCh38 alignment: depth over the variable core relative to the invariant end units and to the unique flanks, the evenness and asymmetry of core depth, mapping quality, soft-clipping and read GC content. A Bayesian ridge regression on standardised features maps them to the diploid length, and VNtyper 2 ships the fitted coefficients as a fixed model file. We fitted on patient exomes only: a depth-ratio model fitted to MucOneUp simulations failed on real exomes, whose repeat reads diverge more from the reference than simulated reads do.

##### 1.12.2 Accuracy

In leave-one-out cross-validation, the estimate was off by 10.8 repeats on average (bootstrap 95% CI 8.8–13.0; $R^2$ 0.66), against 19.2 repeats for predicting the training mean (Supplementary Figure S10a, b). The typical error was about 14 repeats, and the central 95% of errors lay between 29 repeats below and 28 above the PacBio length. Estimates are drawn towards the mean: arrays longer than 130 repeats were underestimated by 12 repeats on average.

##### 1.12.3 Long-array note

In the preregistered simulation, 59dupC sensitivity stayed at 95% or more up to 149 repeats in total and fell to 66% at 150 to 169 repeats, while atypical sensitivity declined steadily, from 87% at 90 to 109 repeats to 49% at 150 to 169 (Supplementary Figure S10c). The note therefore triggers on an estimate above 150 repeats. Because estimates are drawn towards the mean, this cutoff on the estimate caught 7 of the 8 exomes whose true length exceeded 160 repeats (95% CI 53%–98%) and flagged 15 of 76.

##### 1.12.4 Scope

The cutoff was chosen on the same leave-one-out predictions that describe it, so a held-out set has yet to confirm it. Transfer between cohorts was uneven: trained on the German exomes, the model estimated the French ones with a mean error of 10.1 repeats, whereas trained on the French exomes it missed the German ones by 27.1 repeats ($R^2$ -0.69). All training exomes were Twist exomes, so estimates on other kits need local calibration, which the calibration module supports with a laboratory's own PacBio-sized samples (Supplementary Methods S8). The estimate is the sum of both alleles and cannot single out one long allele.

### 2 Supplementary Figures

#### Supplementary Figure S1

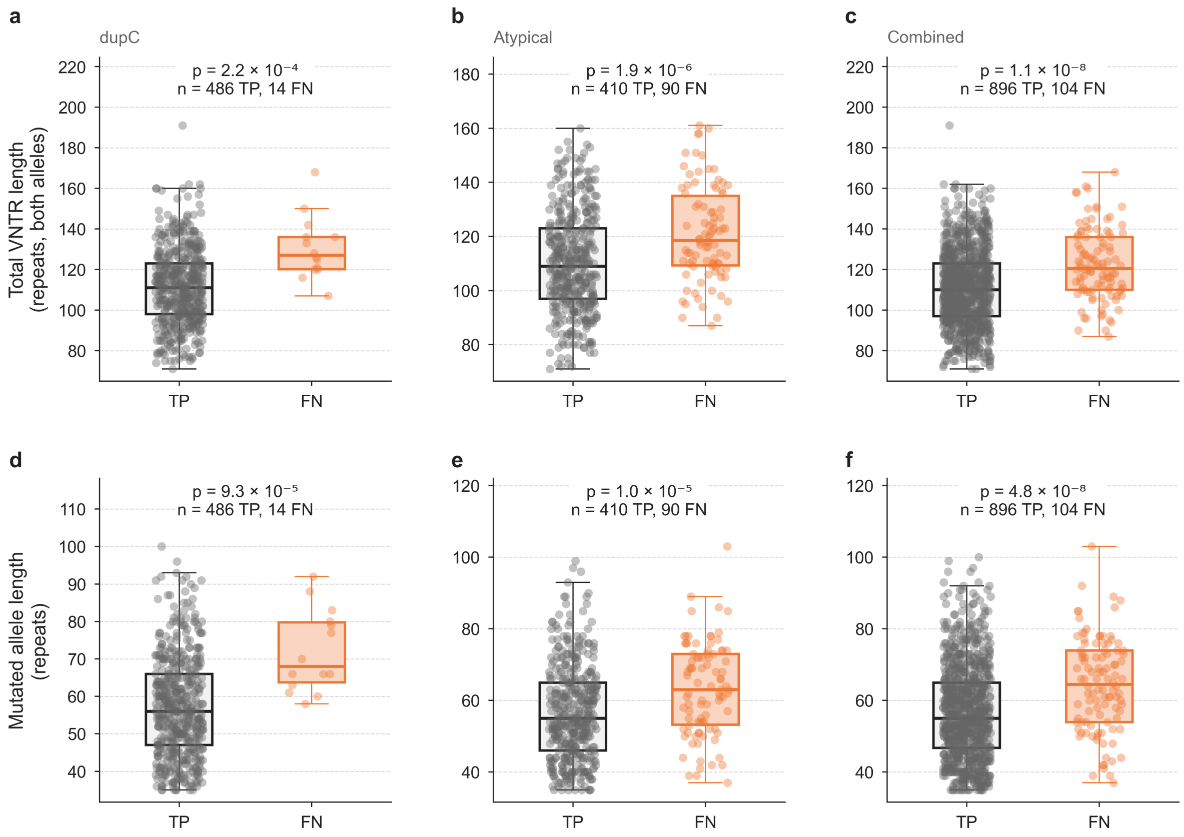

**VNTR length and detection outcome, natural-length simulation.** Total VNTR length of both alleles (top) and mutated-allele length (bottom), in repeats, for true positive (TP, grey) and false negative (FN, orange) samples by variant group (alleles of 35 to 105 repeats). False negatives carried longer alleles in every group (Mann-Whitney U test, p-values shown). Points show samples over box plots. The long-allele simulation is in Supplementary Figure S4.

#### Supplementary Figure S2

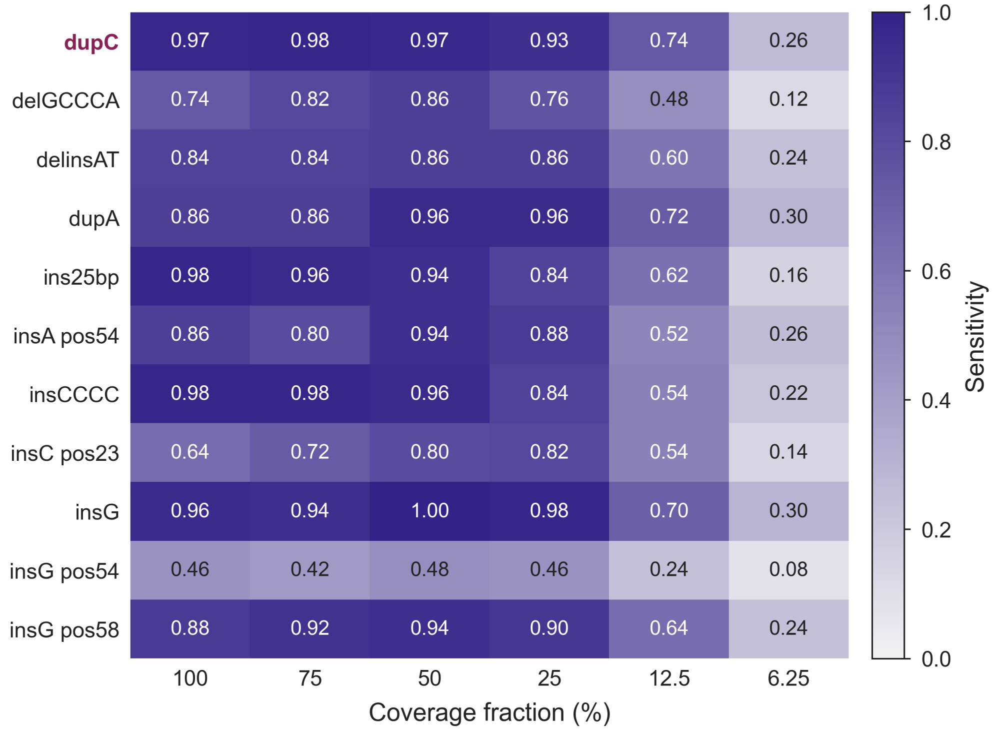

**Mutation-by-coverage sensitivity heatmap, natural-length simulation.** Kestrel sensitivity for each of 11 variant types (rows; 500 59dupC pairs and 50 pairs per atypical type) across six coverage fractions (columns). The first row, labelled in wine, is canonical 59dupC; the other rows are atypical non-dupC frameshifts. Aggregate atypical sensitivity in Figure 2a combines the ten atypical rows and so equals no single row. Specificity was 100.0% at every fraction (Supplementary Table S1c).

#### Supplementary Figure S3

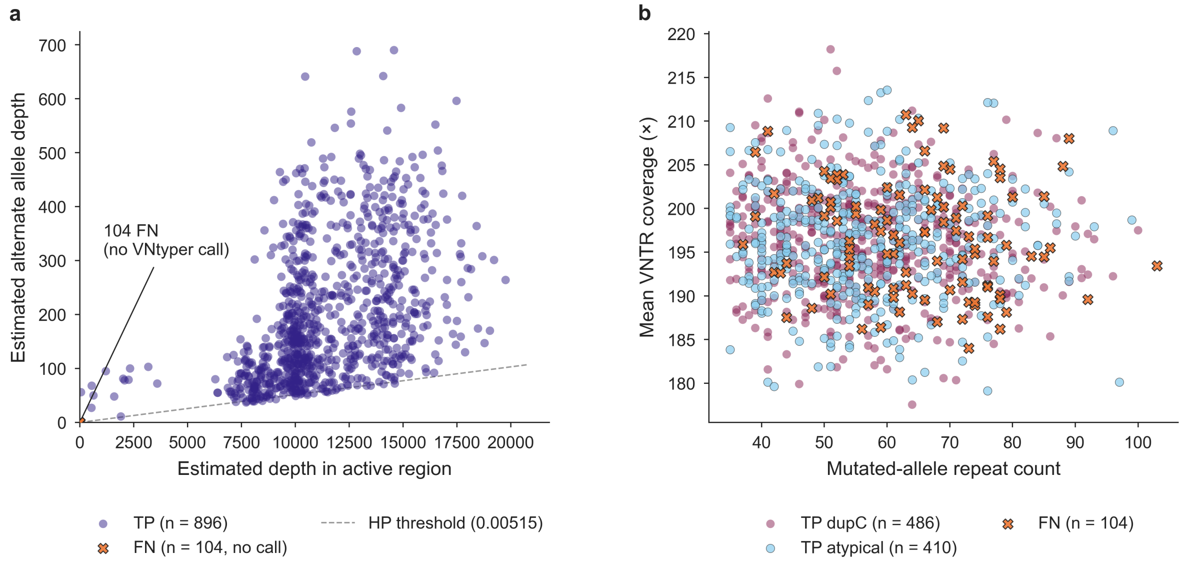

**Simulation depth analysis and false negatives, natural-length simulation.** (**a**) Alternate allele depth versus estimated active VNTR region depth for all 1000 mutated samples. True positives (indigo, n = 896) scale with active-region depth; the dashed line marks the high-precision depth score threshold (0.00515). The 104 false negatives produced no Kestrel call and are shown at the origin. (**b**) Mutated-allele repeat count versus mean VNTR coverage of mutated samples by variant group (59dupC, wine; atypical non-dupC, light blue) and outcome (FN, orange crosses). False negatives had coverage similar to true positives and tended to lie on longer alleles (Supplementary Table S1b); Data S1 lists them per sample.

#### Supplementary Figure S4

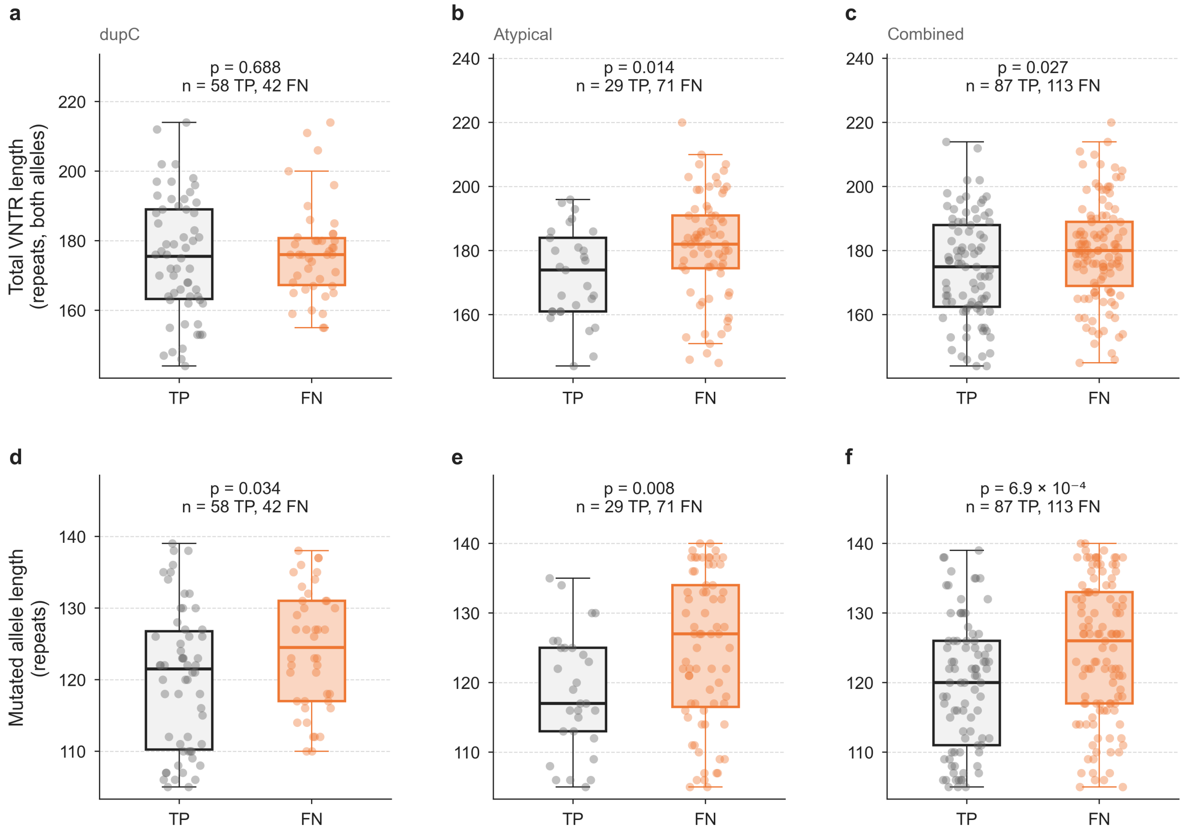

**VNTR length and detection outcome, long-allele simulation.** As Supplementary Figure S1, for 200 pairs whose mutated allele was drawn from 105 to 140 repeats. Kestrel detected 58 of 100 59dupC and 29 of 100 atypical variants, and all 200 wild-type samples were negative.

#### Supplementary Figure S5

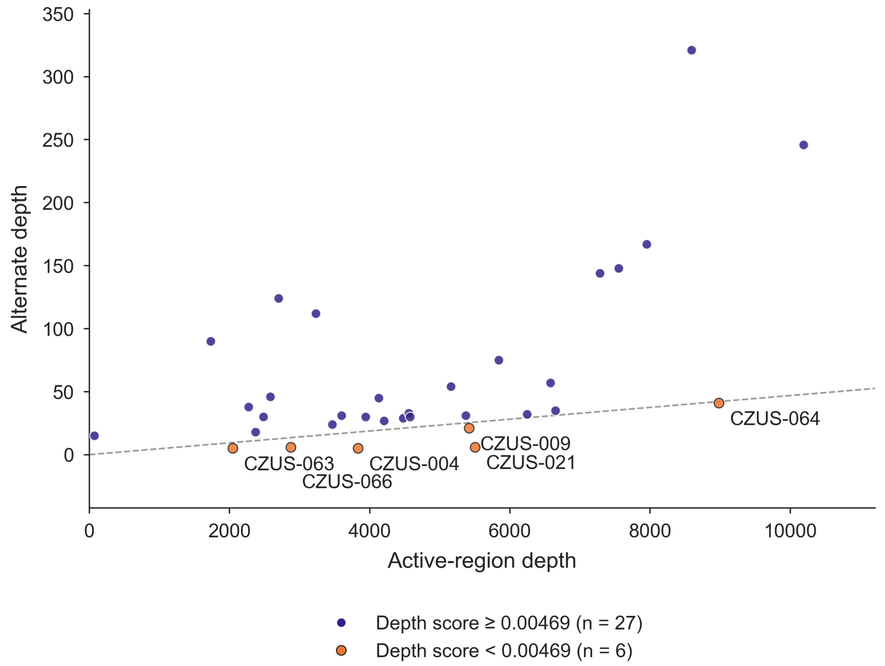

**Depth score analysis in the Czech-US gold standard cohort.** Alternate depth against active-region depth of the best motif/frame-valid, unflagged Kestrel candidate in each of the 33 Czech-US samples that had one (of 83; VNtyper 2.0.47). The line marks the reporting threshold (depth score 0.00469). 27 candidates lay above it, all true positives; 6 lay below it (orange), 5 in PacBio-positive false negatives and 1 in a PacBio-negative sample. The other 2 false negatives had no such candidate. Candidates below the threshold carry display IDs; Supplementary Figure S6 classifies all samples.

#### Supplementary Figure S6

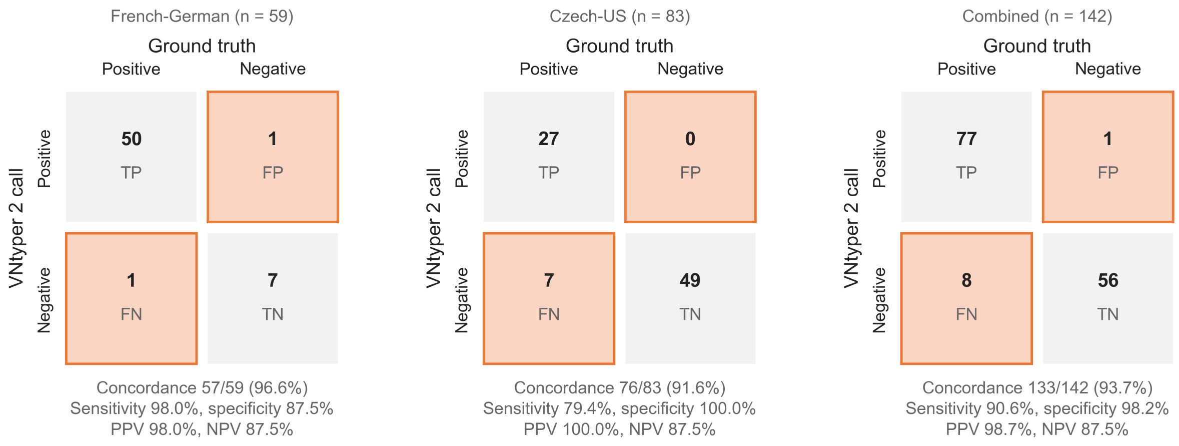

**Per-cohort confusion matrices for gold standard validation.** VNtyper 2 calls against orthogonally confirmed genotypes for the French-German cohort (n=59; SNaPshot and PacBio), the Czech-US cohort (n=83; PacBio) and both combined (n=142). The SNaPshot-only German subset showed 94.7% sensitivity (18/19); French samples were assessed against PacBio-informed ground truth. Grey cells, concordant; orange cells, discordant. TP, true positive; TN, true negative; FP, false positive; FN, false negative; PPV and NPV, positive and negative predictive value.

#### Supplementary Figure S7

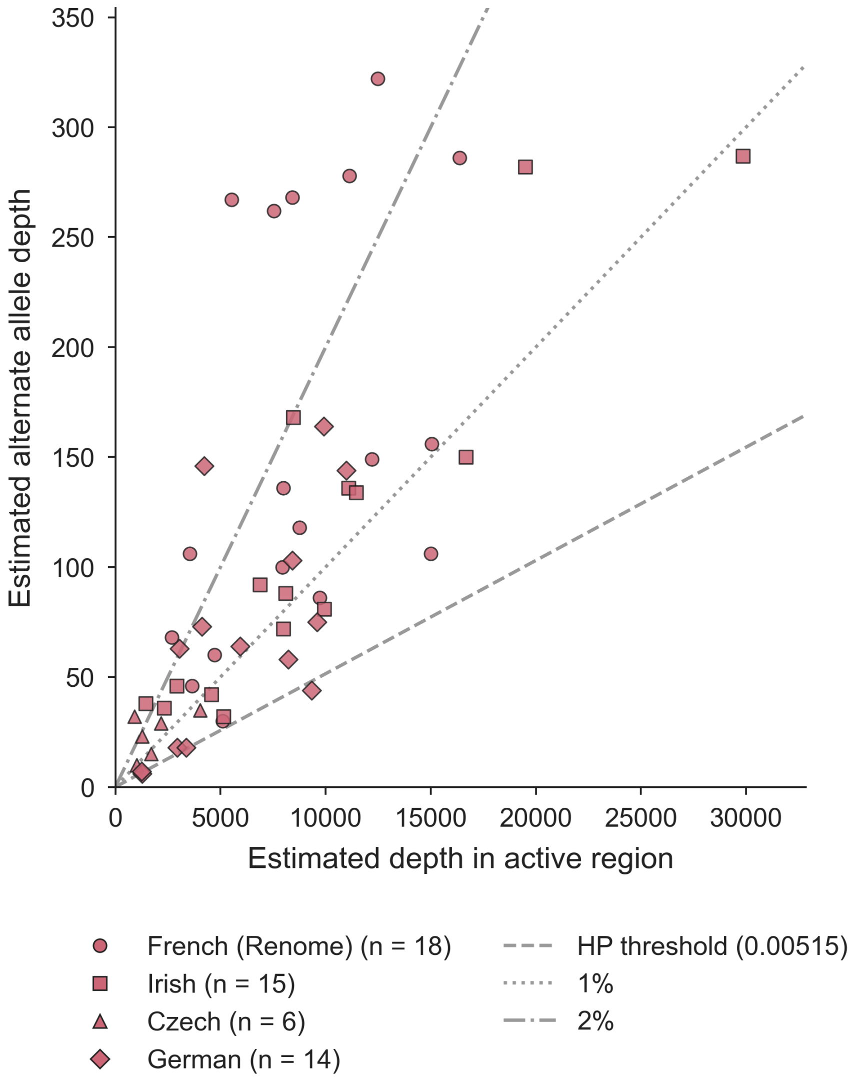

**Depth score distribution in the screening cohorts.** Estimated alternate allele depth versus estimated active VNTR region depth for final positive calls in French Renome (n=18, circles), Irish (n=15, squares), Czech (n=6, triangles) and German (n=14, diamonds) cohorts. Lines mark the high-precision (HP) depth score threshold (0.00515) and reference ratios of 1% and 2%. Most calls lie well above the HP threshold (see Figure 3b). Low-precision (LP) calls from the Czech and German exome cohorts are final calls and are included; the two unconfirmed French targeted-panel LP candidates are not final positives and are not plotted.

### Supplementary Figure S8

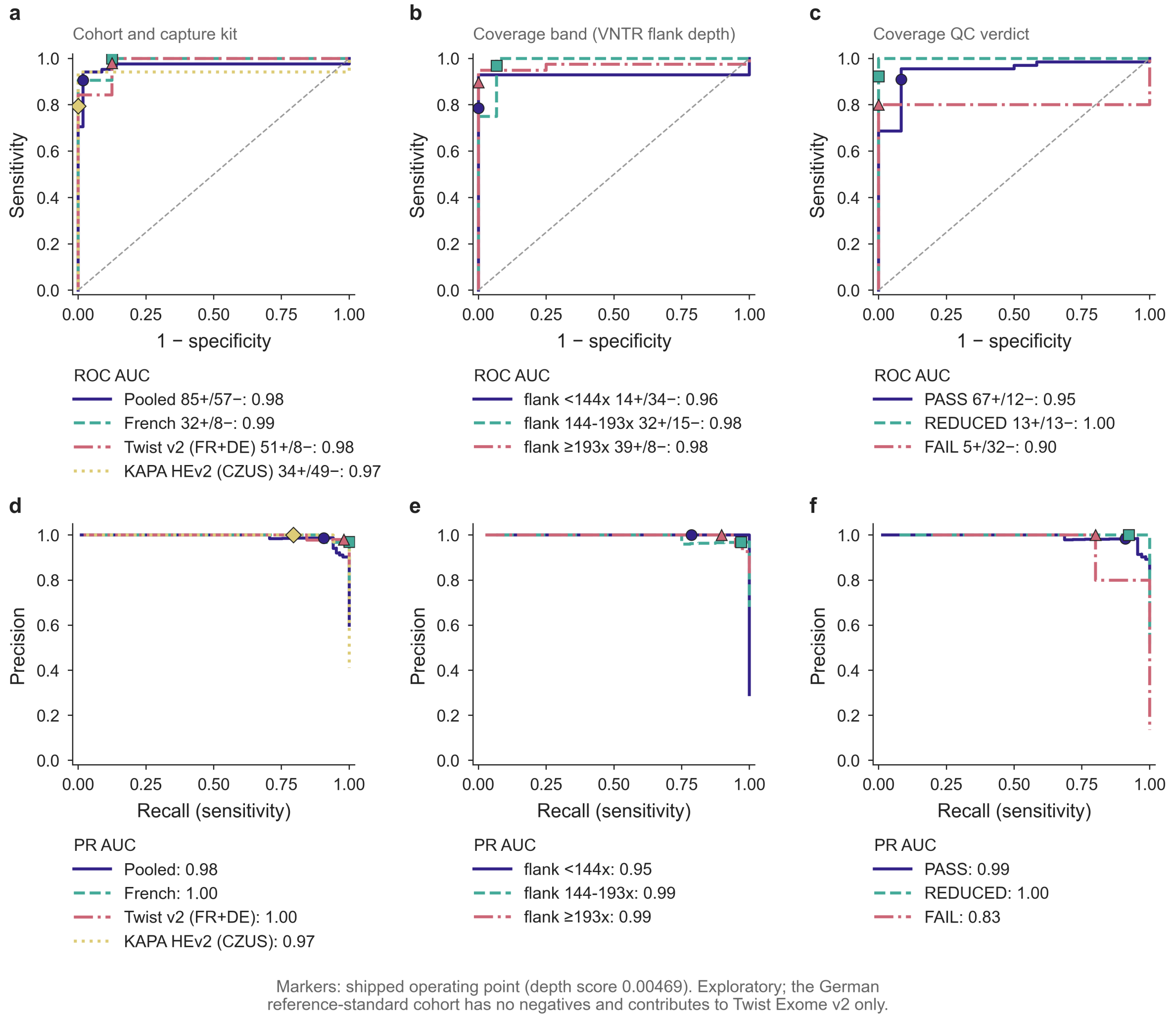

**Exploratory threshold sweep of the VNtyper 2 depth score.** ROC (a-c) and precision-recall (d-f) curves of the sample-level score (Supplementary Methods S8) against the reference standard, by cohort and capture kit (a, d; Twist Exome v2, French and German cohorts; KAPA HyperExome V2, Czech-US cohort), by tertile of mean depth over the unique VNTR flanks (b, e), and by coverage QC verdict (c, f). Markers show the shipped operating point (depth score 0.00469). Legends give positives/negatives and areas under the curve (95% confidence intervals in Supplementary Table S9). The German cohort has no reference negatives and enters only the Twist curve. No threshold was chosen from these curves.

### Supplementary Figure S9

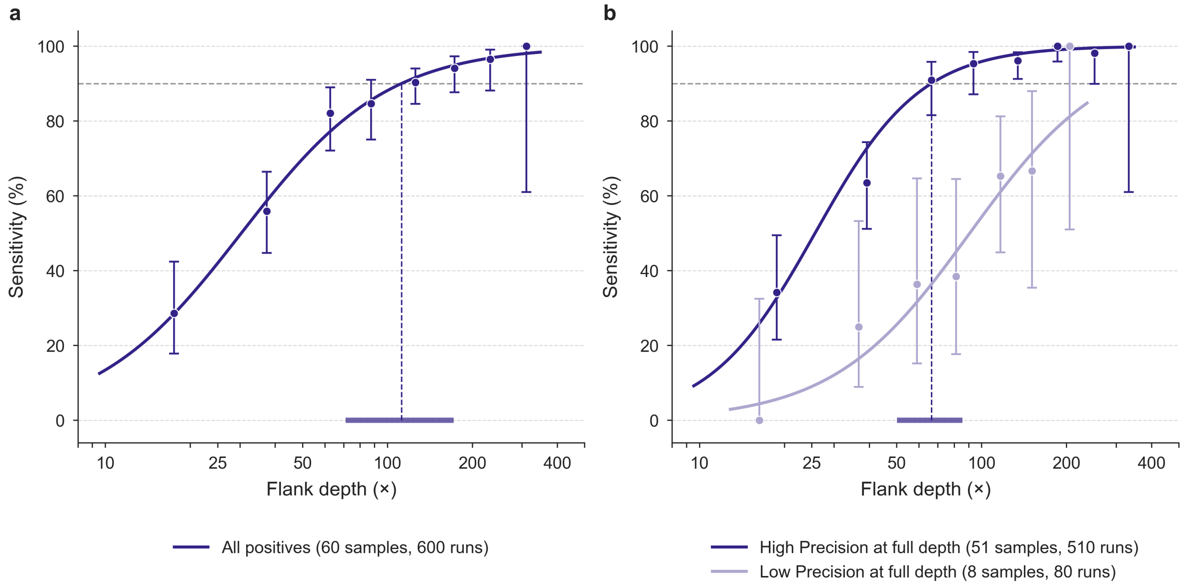

**Coverage-dependent sensitivity in downsampled positive exomes.** Detection of the confirmed variant against mean depth over the unique VNTR flanks in the pooled series (60 confirmed positive exomes, 600 runs, one per sample and fraction; VNtyper 2.0.47; Supplementary Methods S9). Points, observed sensitivity per depth bin (Wilson 95% CI); curves, logistic generalised estimating equations on log2 depth; dashed line and bar above the axis, depth for 90% sensitivity and its cluster-bootstrap 95% CI. (**a**) All positives: 90% sensitivity at 112× (71× to 172×). (**b**) By full-depth confidence tier (post hoc; points offset for legibility): High Precision or High Precision* samples reached 90% at 66× (50× to 85×), whereas the 8 Low Precision samples did not reach 90% within the observed depths. All samples are positive (sensitivity only).

#### Supplementary Figure S10

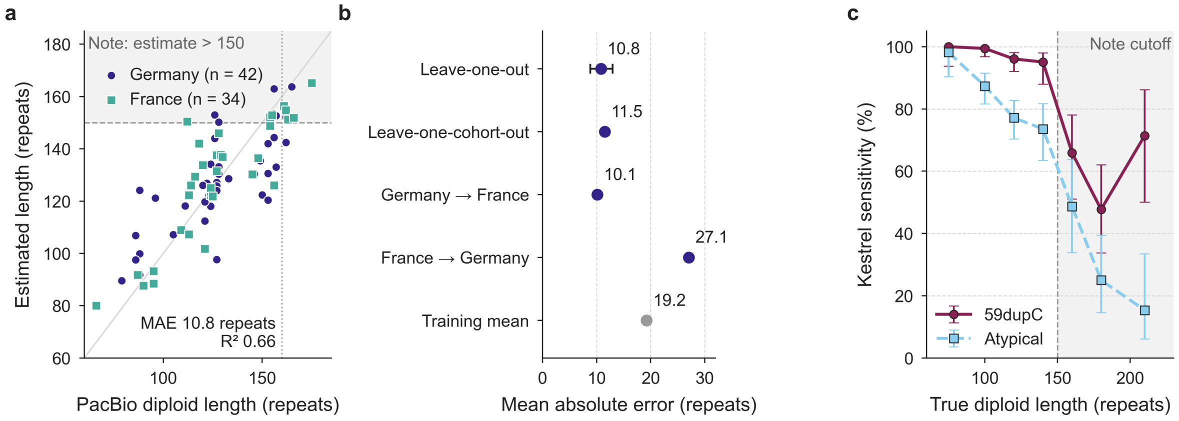

**VNTR length estimate and the long-array note.** (**a**) Estimate from leave-one-out cross-validation against the PacBio diploid length for the 76 Twist exomes the model was trained on (German, circles; French, squares). Grey diagonal, perfect agreement; dashed line and shading, the long-array note at an estimate above 150 repeats; dotted line, a true length of 160 repeats. (**b**) Mean absolute error by validation scheme: leave-one-out with its bootstrap 95% CI, leaving out each of the 5 sub-cohorts in turn, transfer between the German and French exomes in both directions, and predicting the training mean. (**c**) Simulated Kestrel sensitivity of mutated samples by true diploid length in the preregistered natural-length and long-allele simulations, pooled, in bins of 20 repeats (outer bins wider), with Wilson 95% CIs; shading, lengths above the note cutoff. All lengths count every repeat unit, including the nine invariant units of each allele.

### 3 Supplementary Tables

### Supplementary Table S1: Preregistered held-out simulation

#### (a) Sensitivity by variant type, Kestrel, code-adVNTR and either method

| Variant (pairs) | Natural length: Kestrel | Natural length: adVNTR | Natural length: either | Long allele: Kestrel | Long allele: adVNTR | Long allele: either |
|---|---|---|---|---|---|---|
| dupC (500 / 100) | 97.2% (95.4–98.3) | 95.6% (93.4–97.1) | 97.6% (95.9–98.6) | 58.0% (48.2–67.2) | 47.0% (37.5–56.7) | 58.0% (48.2–67.2) |
| Atypical, pooled (500 / 100) | 82.0% (78.4–85.1) | 71.2% (67.1–75.0) | 93.8% (91.3–95.6) | 29.0% (21.0–38.5) | 30.0% (21.9–39.6) | 41.0% (31.9–50.8) |
| delGCCCA (50 / 10) | 74.0% (60.4–84.1) | 30.0% (19.1–43.8) | 82.0% (69.2–90.2) | 30.0% (10.8–60.3) | 20.0% (5.7–51.0) | 40.0% (16.8–68.7) |
| delinsAT (50 / 10) | 84.0% (71.5–91.7) | 92.0% (81.2–96.8) | 92.0% (81.2–96.8) | 20.0% (5.7–51.0) | 40.0% (16.8–68.7) | 40.0% (16.8–68.7) |
| dupA (50 / 10) | 86.0% (73.8–93.0) | 8.0% (3.2–18.8) | 90.0% (78.6–95.7) | 30.0% (10.8–60.3) | 0.0% (0.0–27.8) | 30.0% (10.8–60.3) |
| ins25bp (50 / 10) | 98.0% (89.5–99.6) | 52.0% (38.5–65.2) | 100.0% (92.9–100.0) | 40.0% (16.8–68.7) | 40.0% (16.8–68.7) | 60.0% (31.3–83.2) |
| insA_pos54 (50 / 10) | 86.0% (73.8–93.0) | 98.0% (89.5–99.6) | 98.0% (89.5–99.6) | 40.0% (16.8–68.7) | 40.0% (16.8–68.7) | 40.0% (16.8–68.7) |
| insC_pos23 (50 / 10) | 64.0% (50.1–75.9) | 42.0% (29.4–55.8) | 82.0% (69.2–90.2) | 20.0% (5.7–51.0) | 0.0% (0.0–27.8) | 20.0% (5.7–51.0) |
| insCCCC (50 / 10) | 98.0% (89.5–99.6) | 96.0% (86.5–98.9) | 100.0% (92.9–100.0) | 50.0% (23.7–76.3) | 30.0% (10.8–60.3) | 50.0% (23.7–76.3) |
| insG (50 / 10) | 96.0% (86.5–98.9) | 98.0% (89.5–99.6) | 98.0% (89.5–99.6) | 20.0% (5.7–51.0) | 30.0% (10.8–60.3) | 30.0% (10.8–60.3) |
| insG_pos54 (50 / 10) | 46.0% (33.0–59.6) | 100.0% (92.9–100.0) | 100.0% (92.9–100.0) | 0.0% (0.0–27.8) | 20.0% (5.7–51.0) | 20.0% (5.7–51.0) |
| insG_pos58 (50 / 10) | 88.0% (76.2–94.4) | 96.0% (86.5–98.9) | 96.0% (86.5–98.9) | 40.0% (16.8–68.7) | 80.0% (49.0–94.3) | 80.0% (49.0–94.3) |
| Specificity, wild type (TN/n) | 1000/1000 | 1000/1000 | 1000/1000 | 200/200 | 200/200 | 200/200 |

#### (b) Sensitivity by mutated-allele length

| Simulation, group | Mutated allele (repeats) | Kestrel TP/n | Kestrel | adVNTR | Either |
|---|---|---|---|---|---|
| Natural length, dupC | 35–60 | 302/304 | 99.3% (97.6–99.8) | 99.7% (98.2–99.9) | 99.7% (98.2–99.9) |
| Natural length, dupC | 61–85 | 169/179 | 94.4% (90.0–96.9) | 89.9% (84.7–93.5) | 95.0% (90.7–97.3) |
| Natural length, dupC | 86–105 | 15/17 | 88.2% (65.7–96.7) | 82.4% (59.0–93.8) | 88.2% (65.7–96.7) |
| Natural length, Atypical | 35–60 | 265/303 | 87.5% (83.3–90.7) | 71.9% (66.6–76.7) | 97.0% (94.5–98.4) |
| Natural length, Atypical | 61–85 | 135/184 | 73.4% (66.6–79.2) | 70.7% (63.7–76.8) | 88.6% (83.2–92.4) |
| Natural length, Atypical | 86–105 | 10/13 | 76.9% (49.7–91.8) | 61.5% (35.5–82.3) | 92.3% (66.7–98.6) |
| Long allele, dupC | 105–115 | 20/27 | 74.1% (55.3–86.8) | 63.0% (44.2–78.5) | 74.1% (55.3–86.8) |
| Long allele, dupC | 116–125 | 20/34 | 58.8% (42.2–73.6) | 47.1% (31.5–63.3) | 58.8% (42.2–73.6) |
| Long allele, dupC | 126–140 | 18/39 | 46.2% (31.6–61.4) | 35.9% (22.7–51.6) | 46.2% (31.6–61.4) |
| Long allele, Atypical | 105–115 | 10/26 | 38.5% (22.4–57.5) | 38.5% (22.4–57.5) | 50.0% (32.1–67.9) |
| Long allele, Atypical | 116–125 | 13/32 | 40.6% (25.5–57.7) | 37.5% (22.9–54.7) | 53.1% (36.4–69.1) |
| Long allele, Atypical | 126–140 | 6/42 | 14.3% (6.7–27.8) | 19.0% (10.0–33.3) | 26.2% (15.3–41.1) |

##### (c) Coverage titration, Kestrel, natural-length simulation

| Coverage (%) | dupC sensitivity | Atypical sensitivity | Specificity (TN/n) |
|---|---|---|---|
| 75 | 97.6% (95.9–98.6) | 82.6% (79.0–85.7) | 1000/1000 |
| 50 | 96.8% (94.9–98.0) | 87.4% (84.2–90.0) | 1000/1000 |
| 25 | 93.2% (90.6–95.1) | 83.0% (79.5–86.0) | 1000/1000 |
| 12.5 | 74.0% (70.0–77.7) | 56.0% (51.6–60.3) | 1000/1000 |
| 6.25 | 26.4% (22.7–30.4) | 20.6% (17.3–24.4) | 1000/1000 |

Natural-length simulation, alleles of 35 to 105 repeats; long-allele simulation, mutated allele of 105 to 140 repeats (Supplementary Methods S3). Sensitivity with Wilson 95% confidence intervals. “Either” (exploratory), positive when Kestrel or code-adVNTR made an unflagged call. Specificity, wild-type samples without a positive call. Full coverage for (c) is the natural-length Kestrel column of (a). Only unflagged calls count as positive; 4-bp insertion artifacts never do. No result was missing.

#### Supplementary Table S2: Cohort characteristics

| Cohort | N | Set | Ver. | Capture | Platform | Build | Asc. |
|---|---|---|---|---|---|---|---|
| French | 40 | GS | 2.0.47 | Twist v2 | NVX / NS500 | hg38 / hg19 | SN + PB |
| German | 19 | GS | 2.0.47 | Twist v2 | NV6 | hg38 | SN |
| Czech-US | 83 | GS | 2.0.47 | KAPA HEv2 | NVX | hg19 | PB |
| French (Renome) | 1583 | Screen | 2.0.47 | Twist panel | NS500 / MiSeq | hg19 | Unsel. CKD |
| Czech | 180 | Screen | 2.0.47 | KAPA HEv2 | NA | hg19 | Suspect ADTKD |
| Irish | 777 | Screen | 2.0.47 | Twist+CeGaT | NA | hg38 / hg19 | Unsel. CKD |
| German (CeRKiD) | 1043 | Screen | 2.0.47 | Twist v2 | NV6 / NVX | hg38 | Unsel. CKD |

All cohorts used Illumina short-read sequencing. Set, cohort role; GS, gold standard; Screen, screening cohort; Ver., VNtyper version; Platform, Illumina sequencer (2 × 150 or 2 × 151 bp paired-end reads, 2 × 159 bp for Czech-US, where known); NV6, NovaSeq 6000; NVX, NovaSeq X; NS500, NextSeq 500/550; NA, not available; Build, reference genome build; Asc., ascertainment or reference standard; Unsel., unselected; CKD, chronic kidney disease; ADTKD, autosomal dominant tubulointerstitial kidney disease; SN, SNaPshot minisequencing; PB, PacBio long-read sequencing; KAPA HEv2, KAPA HyperExome V2 (Roche); Twist+CeGaT, mixed Twist Human Core Exome with RefSeq and CeGaT ExomeXtra; CeRKiD, Charité Center for Rare Kidney Diseases. The French gold standard comprises 34 hg38/BWA-MEM exomes sequenced at Labor Berlin and six hg19 DRAGEN/DRAGMAP exomes (Supplementary Methods S4). The French Renome cohort includes only samples sequenced after the Kachmar et al. 2025 publication cutoff. The Irish cohort excludes 227 Roche targeted sequencing samples lacking MUC1 VNTR coverage.

#### Supplementary Table S3: Performance by cohort, capture kit, and depth

##### (a) By cohort and capture kit

| Cohort, capture kit | Assembly | Pos / Neg | Sensitivity, % (95% CI) | Specificity, % (95% CI) | Flank depth, median (IQR), × | QC PASS / REDUCED / FAIL |
|---|---|---|---|---|---|---|
| French, Twist Exome v2 | GRCh38 (34); GRCh37 (6) | 32 / 8 | 100.0 (89.1–100.0) | 87.5 (47.3–99.7) | 179 (164–200) | 34 / 1 / 5 |

| Cohort, capture kit | Assembly | Pos / Neg | Sensitivity, % (95% CI) | Specificity, % (95% CI) | Flank depth, median (IQR), × | QC PASS / REDUCED / FAIL |
|---|---|---|---|---|---|---|
| German, Twist Exome v2 | GRCh38 (19) | 19 / 0 | 94.7 (74.0–99.9) | NA | 268 (233–292) | 16 / 3 / 0 |
| Twist Exome v2, pooled | GRCh38 (53); GRCh37 (6) | 51 / 8 | 98.0 (89.6–100.0) | 87.5 (47.3–99.7) | 198 (168–232) | 50 / 4 / 5 |
| Czech-US, KAPA HyperExome V2 | GRCh37 (83) | 34 / 49 | 79.4 (62.1–91.3) | 100.0 (92.7–100.0) | 141 (79–178) | 29 / 22 / 32 |
| All reference-standard samples | GRCh37 (89); GRCh38 (53) | 85 / 57 | 90.6 (82.3–95.8) | 98.2 (90.6–100.0) | 168 (124–208) | 79 / 26 / 37 |

#### (b) Sensitivity by stratum

| Samples | Stratum | Level | Detected / positives | Sensitivity, % (95% CI) |
|---|---|---|---|---|
| All | Flank depth | <50× | 0 / 1 | 0.0 (0.0–97.5) |
| All | Flank depth | 50–<100× | 1 / 2 | 50.0 (1.3–98.7) |
| All | Flank depth | ≥100× | 76 / 82 | 92.7 (84.8–97.3) |
| All | Coverage QC | PASS | 61 / 67 | 91.0 (81.5–96.6) |
| All | Coverage QC | REDUCED | 12 / 13 | 92.3 (64.0–99.8) |
| All | Coverage QC | FAIL | 4 / 5 | 80.0 (28.4–99.5) |
| All | Assembly | GRCh37 | 32 / 39 | 82.1 (66.5–92.5) |
| All | Assembly | GRCh38 | 45 / 46 | 97.8 (88.5–99.9) |
| All | Sequencing platform | Illumina NextSeq 500/550 | 5 / 5 | 100.0 (47.8–100.0) |
| All | Sequencing platform | Illumina NovaSeq 6000 | 18 / 19 | 94.7 (74.0–99.9) |
| All | Sequencing platform | Illumina NovaSeq X | 27 / 27 | 100.0 (87.2–100.0) |
| All | Sequencing platform | Illumina NovaSeq X Plus | 27 / 34 | 79.4 (62.1–91.3) |
| All | Reference variant class | 59dupC | 66 / 70 | 94.3 (86.0–98.4) |
| All | Reference variant class | atypical | 11 / 15 | 73.3 (44.9–92.2) |
| Czech-US | Flank depth | <50× | 0 / 1 | 0.0 (0.0–97.5) |
| Czech-US | Flank depth | 50–<100× | 1 / 2 | 50.0 (1.3–98.7) |
| Czech-US | Flank depth | ≥100× | 26 / 31 | 83.9 (66.3–94.5) |
| Czech-US | Coverage QC | PASS | 19 / 24 | 79.2 (57.8–92.9) |
| Czech-US | Coverage QC | REDUCED | 8 / 9 | 88.9 (51.8–99.7) |
| Czech-US | Coverage QC | FAIL | 0 / 1 | 0.0 (0.0–97.5) |

| Samples | Stratum | Level | Detected / positives | Sensitivity, % (95% CI) |
|---|---|---|---|---|
| Czech-US | Assembly | GRCh37 | 27 / 34 | 79.4 (62.1–91.3) |
| Czech-US | Sequencing platform | Illumina NovaSeq X Plus | 27 / 34 | 79.4 (62.1–91.3) |
| Czech-US | Reference variant class | 59dupC | 16 / 19 | 84.2 (60.4–96.6) |
| Czech-US | Reference variant class | atypical | 11 / 15 | 73.3 (44.9–92.2) |
| All | Mutated allele (PacBio repeats) | <60 | 13 / 14 | 92.9 (66.1–99.8) |
| All | Mutated allele (PacBio repeats) | 60–79 | 16 / 19 | 84.2 (60.4–96.6) |
| All | Mutated allele (PacBio repeats) | ≥80 | 21 / 25 | 84.0 (63.9–95.5) |
| Czech-US | Mutated allele (PacBio repeats) | <60 | 10 / 11 | 90.9 (58.7–99.8) |
| Czech-US | Mutated allele (PacBio repeats) | 60–79 | 10 / 13 | 76.9 (46.2–95.0) |
| Czech-US | Mutated allele (PacBio repeats) | ≥80 | 7 / 10 | 70.0 (34.8–93.3) |

##### (c) Firth-penalised logistic models of detection among reference positives

| Model | n (missed) | Term | Odds ratio (95% CI) | P |
|---|---|---|---|---|
| M1 | 85 (8) | French / Twist Exome v2 vs Czech-US / KAPA | 17.73 (2.00–2339) | 0.005 |
| M1 | 85 (8) | German / Twist Exome v2 vs Czech-US / KAPA | 3.36 (0.65–33.82) | 0.158 |
| M2 | 85 (8) | French / Twist Exome v2 vs Czech-US / KAPA | 16.54 (1.82–2191) | 0.008 |
| M2 | 85 (8) | German / Twist Exome v2 vs Czech-US / KAPA | 2.19 (0.33–24.38) | 0.430 |
| M2 | 85 (8) | log2(flank depth / 100×), per doubling | 2.07 (0.61–8.06) | 0.241 |
| M3 (Czech-US only) | 34 (7) | log2(flank depth / 100×), per doubling | 1.96 (0.56–7.78) | 0.288 |

VNtyper 2.0.47 on the reference-standard samples (Supplementary Methods S10). Pos / Neg, reference-positive / reference-negative samples; flank depth, mean depth over the unique VNTR flanks; QC, coverage verdict (build-specific limits on the VNTR window mean). Proportions have exact (Clopper-Pearson) intervals; odds ratios have profile penalised-likelihood intervals. The cohort-and-kit term compares each Twist cohort with Czech-US, KAPA HyperExome V2. Few positives lie below 100× flank depth, so the low-depth bands are descriptive. Only unflagged calls count as positive; 4-bp insertion artifacts never do.

#### Supplementary Table S4: Variant stratification by cohort

| Variant | Call | French | Irish | Czech | German | Total |
|---|---|---|---|---|---|---|
| 59dupC | Final, n (HP/HP*/LP) | 15 (4/11/0) | 14 (8/6/0) | 3 (1/0/2) | 12 (3/4/5) | 44 |
| 59dupC | Candidate LP | 2 | 0 | 0 | 0 | 2 |
| 59dupC | Flagged | 0 | 0 | 0 | 0 | 0 |
| Atyp. | Final, n (HP/HP*/LP) | 3 (1/2/0) | 1 (1/0/0) | 3 (3/0/0) | 2 (2/0/0) | 9 |
| Atyp. | Candidate LP | 0 | 0 | 0 | 0 | 0 |
| Atyp. | Flagged, n (AF/LD) | 0 (0/0) | 0 (0/0) | 1 (0/1) | 0 (0/0) | 1 (0/1) |
| Total | Final positives | 18 | 15 | 6 | 14 | 53 |
| Total | Candidate LP | 2 | 0 | 0 | 0 | 2 |

59dupC, the VNtyper 2 single-base insertion at position 67; Atyp., atypical non-dupC frameshift variant. Final calls passed all quality filters and cohort-specific interpretation rules (Supplementary Methods S6); the French targeted-panel LP calls are candidates, not final positives. AF, artifact-filtered 4-bp insertion; LD, low-depth call in a conserved motif; HP, High Precision; HP*, High Precision*; LP, Low Precision. PacBio confirmed atypical non-dupC variants in the Czech-US validation samples; atypical screening calls outside those samples require orthogonal confirmation.

#### Supplementary Table S5: Screening detection-rate meta-analysis

| Cohort | Final positives | N | Detection rate, % |
|---|---|---|---|
| French Renome | 18 | 1583 | 1.14 |
| Irish | 15 | 777 | 1.93 |
| German | 14 | 1043 | 1.34 |
| Random-effects pooled | 47 | 3403 | 1.42 (0.72–2.79) |

Logit-scale random-effects summary across the French Renome, Irish and German unselected chronic kidney disease cohorts. 95% prediction interval 0.6%–3.2%; $Q = 2.38$, $P = 0.30$; $I^2 = 16.0\%$; $\tau^2 = 0.012$.

#### Supplementary Table S6: VNtyper 1.3.0 versus VNtyper 2.0.47 on the reference-standard cohorts

| Cohort (n) | Version | TP | FN | TN | FP | 4-bp AF | Sens. (95% CI) | Spec. (95% CI) |
|---|---|---|---|---|---|---|---|---|
| French (40) | 1.3.0, 4-bp rule | 32 | 0 | 7 | 1 | 1 | 100.0% (89.3–100.0) | 87.5% (52.9–97.8) |
| French (40) | 1.3.0, no 4-bp rule | 32 | 0 | 6 | 2 | 0 | 100.0% (89.3–100.0) | 75.0% (40.9–92.9) |
| French (40) | 2.0.47 | 32 | 0 | 7 | 1 | 0 | 100.0% (89.3–100.0) | 87.5% (52.9–97.8) |
| German (19) | 1.3.0, 4-bp rule | 18 | 1 | 0 | 0 | 0 | 94.7% (75.4–99.1) | NA |
| German (19) | 1.3.0, no 4-bp rule | 18 | 1 | 0 | 0 | 0 | 94.7% (75.4–99.1) | NA |
| German (19) | 2.0.47 | 18 | 1 | 0 | 0 | 0 | 94.7% (75.4–99.1) | NA |
| Czech-US (83) | 1.3.0, 4-bp rule | 27 | 7 | 49 | 0 | 0 | 79.4% (63.2–89.7) | 100.0% (92.7–100.0) |
| Czech-US (83) | 1.3.0, no 4-bp rule | 27 | 7 | 49 | 0 | 0 | 79.4% (63.2–89.7) | 100.0% (92.7–100.0) |
| Czech-US (83) | 2.0.47 | 27 | 7 | 49 | 0 | 0 | 79.4% (63.2–89.7) | 100.0% (92.7–100.0) |
| Pooled (142) | 1.3.0, 4-bp rule | 77 | 8 | 56 | 1 | 1 | 90.6% (82.5–95.2) | 98.2% (90.7–99.7) |
| Pooled (142) | 1.3.0, no 4-bp rule | 77 | 8 | 55 | 2 | 0 | 90.6% (82.5–95.2) | 96.5% (88.1–99.0) |
| Pooled (142) | 2.0.47 | 77 | 8 | 56 | 1 | 0 | 90.6% (82.5–95.2) | 98.2% (90.7–99.7) |

Kestrel calls from the same BAM files, counted as in Supplementary Methods S7. “4-bp rule” applies the VNtyper 2 4-bp insertion artifact filter to VNtyper 1.3.0 output; “no 4-bp rule” shows VNtyper 1.3.0 as released. Specificity is undefined for the German cohort, which has no reference negatives. The French false positive in all rows is the discordant, unresolved sample FR-GS-006. Sens., sensitivity; Spec., specificity; TP, true positive; FN, false negative; TN, true negative; FP, false positive; 4-bp AF, samples whose only call was a filtered 4-bp insertion artifact. Wilson score 95% confidence intervals.

#### Supplementary Table S7: Differences between VNtyper 1.3.0 and VNtyper 2.0.47

| Property | VNtyper 1.3.0 | VNtyper 2.0.47 |
|---|---|---|
| 4-bp insertion artifacts | Reported as Low Precision calls (5 rows) | Flagged and filtered automatically; never positive |
| Variant scope | 1-bp insertions (including 59dupC), multi-base insertions, deletions; all passing Kestrel rows listed (103 rows) | Same classes; one selected call per sample (84 calls) with VNTR variant nomenclature |

| Property | VNtyper 1.3.0 | VNtyper 2.0.47 |
|---|---|---|
| Runtime, Kestrel only (matched) | Median 9.1 s | Median 11.8 s; 9.8 s in fast mode |
| Runtime, Kestrel and code-adVNTR (matched) | Median 56 min on one core (n = 5) | Median 30.5 s; 107-fold faster |
| Runtime in cohort analyses | Not recorded per sample | Median 124.2 s (French), 153.6 s (German), 146.4 s (Czech) per exome and 40.8 s per panel with code-adVNTR under shared load; Irish exomes 98.5 s at 4 threads (12.9 s fast mode) |
| Scalability | Per-sample output only | Cohort mode; 910 samples per hour with four concurrent runs; 6351 panels in one job (8.6 h) |
| Flagging | None | Configurable rules (4-bp insertion artifact; low depth in conserved motifs) |
| Confidence tiers | High Precision, High Precision*, Low Precision | Same tiers plus an explicit Negative tier for subthreshold candidates |
| Coverage QC | None | VNTR and flank depth, uncovered fraction, build-specific PASS/REDUCED/FAIL verdict |
| code-adVNTR | Optional but slow, so usually skipped; not run in the accuracy comparison | Maintained fork with compiled Viterbi core (median 18.2 s per sample at 8 threads); default on VNtyper-Online, cross-matched with Kestrel |
| Inputs | BAM, FASTQ | BAM, CRAM, FASTQ; automatic genome build and chromosome-naming detection |
| Robustness to input | code-adVNTR aborts silently on region slices with mates outside the slice (20 of 20 benchmark slices) | Runs on region slices, including reads without their mate |
| code-adVNTR on GRCh38 input | Realigns to a user-supplied GRCh37 chromosome 1 reference | Native GRCh38 model of the full VNTR array |
| Additional methods | None | SHARK read prefilter with recorded provenance; VNTR length estimate with long-array note; local calibration on a laboratory’s confirmed samples |
| Variant names | Kestrel motif coordinates (for example 67:G>GG) | VNTR nomenclature on the canonical repeat unit (for example 59dupC), with naming tier |
| Outputs | Per-sample variant TSV | HTML report with an interactive IGV.js read view, variant name, confidence grade and coverage verdict; JSON summary; cohort tables |
| Web access | None | VNtyper-Online with browser-side region extraction and a REST API |

| Property | VNtyper 1.3.0 | VNtyper 2.0.47 |
|---|---|---|
| Deployment | Unpinned installation script | Versioned PyPI package and non-root container image pinned by digest |
| Software testing | None | Unit, integration and reference-output tests in continuous integration; 91% line and branch coverage floor |

Accuracy and sample-status agreement are in Supplementary Table S6; matched runtimes in Supplementary Methods S7. QC, quality control.

#### Supplementary Table S8: Discordant cases in the reference-standard cohorts

| Case | Reference | Reference result | VNtyper 2 call | Flank depth, QC | Resolved | Counted as | Remaining uncertainty |
|---|---|---|---|---|---|---|---|
| CZUS-004 | PacBio | 54_55insG | Negative | 280×, PASS | yes | FN | None for carrier status (allele lengths in Table S10) |
| CZUS-009 | PacBio | 58_59insG | Negative | 174×, PASS | yes | FN | None for carrier status (allele lengths in Table S10) |
| CZUS-015 | PacBio | 23dupC | Negative | 265×, PASS | yes | FN | None for carrier status (allele lengths in Table S10) |
| CZUS-040 | PacBio | 59dupC | Negative | 47×, FAIL | yes | FN | None for carrier status (allele lengths in Table S10) |
| CZUS-063 | PacBio | 59dupC | Negative | 84×, REDUCED | yes | FN | None for carrier status (allele lengths in Table S10) |
| CZUS-064 | PacBio | 23dupC | Negative | 220×, PASS | yes | FN | None for carrier status (allele lengths in Table S10) |
| CZUS-066 | PacBio | 59dupC | Negative | 124×, PASS | yes | FN | None for carrier status (allele lengths in Table S10) |
| DE-GS-015 | SNaPshot | 59dupC | Negative | 218×, PASS | yes (dupC) | FN | SNaPshot tests 59dupC only |
| FR-GS-006 | PacBio | negative | 67:G>GG (High Precision) | 169×, PASS | no | FP (primary analysis) | Called by VNtyper 1.3.0, VNtyper 2 and code-adVNTR; SNaPshot uninterpretable; PacBio negative |

| Case | Reference | Reference result | VNtyper 2 call | Flank depth, QC | Resolved | Counted as | Remaining uncertainty |
|---|---|---|---|---|---|---|---|
| CZUS-022 | PacBio | 58_59insG | 67:G>GG (High Precision) | 277×, PASS | yes (same variant) | TP (carrier level) | Same variant: VNtyper 2 names it 58_59insG; Kestrel records the insertion at the dupC position |
| CZUS-023 | PacBio | del14b | 28:C>C+46 bp ins (High Precision) | 267×, PASS | yes (allele differs) | TP (carrier level) | Carrier status concordant; PacBio 14-bp deletion, VNtyper 2 +46-bp insertion, both net +1 frameshifts |
| CZUS-045 | PacBio | 60dupA | 61:T>TT (High Precision) | 176×, PASS | yes (same variant) | TP (carrier level) | Same variant: VNtyper 2 names it 60dupA |
| CZUS-057 | PacBio | 60dupA | 22:G>G+16 bp ins (High Precision) | 140×, REDUCED | yes (allele differs) | TP (carrier level) | Carrier status concordant; PacBio 60dupA, VNtyper 2 a multi-base insertion at another position |
| FR-GS-035 | PacBio | positive (type not reported) | 60:C>CT (High Precision) | 179×, FAIL | partly | TP (carrier level) | Carrier status concordant; the reference reported carrier status only, so the variant type is unresolved |

Every reference-standard sample in which VNtyper 2.0.47 disagreed with the reference: 8 false negatives, 1 false positive, and 5 carriers detected with a variant written differently from the reference (true positives at carrier level). The PacBio variant table showed the same variant in CZUS-022 and CZUS-045 and a different allele with the same carrier status in CZUS-023 and CZUS-057. FR-GS-006 was negative by PacBio and counts as a false positive in the primary analysis; French specificity is 100.0% both if it is excluded (7/7; 95% CI 64.6%–100.0%) and if it is counted as a true negative (8/8; 95% CI 67.6%–100.0%). Flank depth, mean depth over the unique VNTR flanks; QC, coverage QC verdict; FN, false negative; FP, false positive; TP, true positive.

### Supplementary Table S9: Exploratory threshold sweep of the depth score

#### a. Area under the curve and robustness range by stratum.

| Stratum | Pos/Neg | ROC AUC (95% CI) | PR AUC (95% CI) | Sens. / Spec. at 0.00469, % | Unchanged range | Within 5 points |
|---|---|---|---|---|---|---|
| All reference-standard samples | 85/57 | 0.978 (0.953–0.997) | 0.985 (0.967–0.998) | 90.6 / 98.2 | 0.0046–0.0051 | 0.0036–0.0057 |
| Pooled, FR-GS-006 excluded | 85/56 | 0.983 (0.962–0.999) | 0.988 (0.975–0.999) | 90.6 / 100.0 | 0.0046–0.0051 | 0.0036–0.0057 |
| French (Twist v2) | 32/8 | 0.988 (0.953–1.000) | 0.997 (0.988–1.000) | 100.0 / 87.5 | 0.0039–0.0057 | 0.0039–0.0062 |
| Czech-US (KAPA HEv2) | 34/49 | 0.970 (0.925–1.000) | 0.965 (0.913–1.000) | 79.4 / 100.0 | 0.0046–0.0051 | 0.0039–0.0052 |
| Twist v2 (French and German) | 51/8 | 0.980 (0.934–1.000) | 0.997 (0.989–1.000) | 98.0 / 87.5 | 0.0041–0.0052 | 0.0039–0.0057 |
| Flank depth <144× | 14/34 | 0.964 (0.893–1.000) | 0.949 (0.848–1.000) | 78.6 / 100.0 | 0.0025–0.0064 | 0.0025–0.0064 |
| Flank depth 144–193× | 32/15 | 0.983 (0.938–1.000) | 0.991 (0.969–1.000) | 96.9 / 93.3 | 0.0039–0.0057 | 0.0039–0.0057 |
| Flank depth ≥193× | 39/8 | 0.976 (0.929–1.000) | 0.994 (0.984–1.000) | 89.7 / 100.0 | 0.0046–0.0051 | 0.0041–0.0052 |
| Coverage QC PASS | 67/12 | 0.949 (0.878–0.994) | 0.990 (0.977–0.999) | 91.0 / 91.7 | 0.0046–0.0051 | 0.0036–0.0054 |
| Coverage QC REDUCED | 13/13 | 1.000 (1.000–1.000) | 1.000 (1.000–1.000) | 92.3 / 100.0 | 0.0025–0.0057 | 0.0025–0.0057 |
| Coverage QC FAIL | 5/32 | 0.897 (0.691–1.000) | 0.827 (0.481–1.000) | 80.0 / 100.0 | 0.0039–0.0100 | 0.0020–0.0100 |

#### b. Sensitivity and specificity at selected depth-score thresholds.

| Depth-score threshold | Pooled: Sens. / Spec., % | Twist v2: Sens. / Spec., % | KAPA HEv2: Sens. / Spec., % |
|---|---|---|---|
| 0.002 | 96.5 / 87.7 | 100.0 / 12.5 | 91.2 / 100.0 |
| 0.003 | 94.1 / 93.0 | 100.0 / 50.0 | 85.3 / 100.0 |
| 0.0036 | 94.1 / 96.5 | 100.0 / 75.0 | 85.3 / 100.0 |
| 0.004 | 92.9 / 98.2 | 100.0 / 87.5 | 82.4 / 100.0 |
| 0.00469 (shipped) | 90.6 / 98.2 | 98.0 / 87.5 | 79.4 / 100.0 |
| 0.0057 | 85.9 / 98.2 | 94.1 / 87.5 | 73.5 / 100.0 |
| 0.007 | 74.1 / 98.2 | 84.3 / 87.5 | 58.8 / 100.0 |
| 0.008 | 69.4 / 100.0 | 82.4 / 100.0 | 50.0 / 100.0 |
| 0.01 | 57.6 / 100.0 | 66.7 / 100.0 | 44.1 / 100.0 |

Score, replay and counting rules as in Supplementary Methods S8 (FR-GS-006 a false positive except where stated).

Pos/Neg, reference-positive and reference-negative samples. Confidence intervals are class-stratified bootstrap 95% intervals (2000 resamples); intervals truncated at 1.000 reflect small strata, not proven separation. Unchanged range, contiguous threshold interval around 0.00469 in which sensitivity and specificity both equal their shipped values; within 5 points, interval in which each stays within 5 percentage points of its shipped value. Ranges ending at 0.0020 or 0.0100 are cut off by the grid edge. Flank-depth bands are pooled tertiles. The German cohort, without reference negatives, enters only the pooled and Twist v2 rows. Twist v2, Twist Exome v2; KAPA HEv2, KAPA HyperExome V2; PR, precision-recall; Sens., sensitivity; Spec., specificity.

#### Supplementary Table S10: False negatives in the reference-standard cohorts

| Case | Cohort, reference | Truth variant | PacBio repeats (mutated) | Flank / mean cov. (×) | Uncov. % | Coverage QC | Retained cand. | Final DS | Raw DS | Alt / region depth | Subthr. signal | Tier | Retest rec. |
|---|---|---|---|---|---|---|---|---|---|---|---|---|---|
| CZUS-004 | Czech-US, PacBio | 54_55insG | 44 / 63 (long) | 280 / 493 | 0.0 | PASS | yes | 0.00131 | 0.665 | 5 / 3831 | no | Negative | no |
| CZUS-009 | Czech-US, PacBio | 58_59insG | 83 / 95 (long) | 174 / 501 | 0.0 | PASS | yes | 0.00388 | 0.695 | 21 / 5417 | yes | Negative | yes |
| CZUS-015 | Czech-US, PacBio | 23dupC | 44 / 44 (equal) | 265 / 407 | 0.0 | PASS | no | none | 0.678 | none | no | Negative | no |
| CZUS-040 | Czech-US, PacBio | 59dupC | 78 / 79 (short) | 47 / 134 | 0.0 | FAIL | no | none | 0.646 | none | no | Negative | yes |
| CZUS-063 | Czech-US, PacBio | 59dupC | 69 / 82 (long) | 84 / 218 | 0.0 | REDUCED | yes | 0.00244 | 0.631 | 5 / 2046 | yes | Negative | yes |
| CZUS-064 | Czech-US, PacBio | 23dupC | 44 / 80 (long) | 220 / 435 | 0.0 | PASS | yes | 0.00457 | 0.675 | 41 / 8980 | yes | Negative | yes |
| CZUS-066 | Czech-US, PacBio | 59dupC | 44 / 78 (long) | 124 / 303 | 0.0 | PASS | yes | 0.00209 | 0.688 | 6 / 2874 | no | Negative | no |
| DE-GS-015 | German, SNaPshot | 59dupC | 77 / 80 (long) | 218 / 119 | 10.6 | PASS | yes | 0.00409 | 0.664 | 10 / 2445 | no | Negative | no |

All 8 reference positives called negative by VNtyper 2.0.47, all also missed by VNtyper 1.3.0 (Supplementary Table S6). PacBio repeats, short / long allele in repeat units, with the allele carrying the variant (equal, both alleles of the same length); Flank, mean depth over the unique VNTR flanks; mean cov., mean depth over the active VNTR window (build-dependent); Uncov., uncovered active VNTR bases; Retained cand., a motif/frame-valid, unflagged, non-artifact candidate remained after filtering; Final DS, depth score (DS) of the best such candidate (reporting floor 0.00469); Alt / region depth, its alternate k-mer path and active-region depths; Raw DS, highest DS of any Kestrel candidate before frame, motif and flag filters (not callable); Subthr. signal, the pipeline's own rule reported a pathogenic-frame candidate below the floor, so it can differ from Retained cand.; Tier, sample confidence; Retest rec., repeat or orthogonal testing recommended under the proposed testing workflow (coverage QC REDUCED or FAIL, or subthreshold signal).

#### Supplementary Table S11: Coverage-dependent sensitivity in downsampled positive exomes

| Stratum (samples; detected/runs) | Depth | <50× | 50–100× | ≥100× | Depth for 90% (95% CI) | Depth for 95% (95% CI) |
|---|---|---|---|---|---|---|
| All samples, pooled (60; 482/600) | Flank | 57/126 (45%) | 130/156 (83%) | 295/318 (93%) | 112× (71–172) | 175× (101–302) |
| All samples, pooled (60; 482/600) | Window | 72/157 (46%) | 141/164 (86%) | 269/279 (96%) | 85× (58–117) | 122× (77–176) |
| Full-depth High Precision (51; 445/510) | Flank | 54/104 (52%) | 121/130 (93%) | 270/276 (98%) | 66× (50–85) | 92× (65–124) |
| Full-depth High Precision (51; 445/510) | Window | 64/117 (55%) | 122/130 (94%) | 259/263 (98%) | 61× (46–80) | 84× (59–113) |
| Full-depth Low Precision (8; 37/80) | Flank | 3/20 (15%) | 9/24 (38%) | 25/36 (69%) | 308× (122–688)[a] | 468× (157–1201)[a] |
| Full-depth Low Precision (8; 37/80) | Window | 8/36 (22%) | 19/30 (63%) | 10/14 (71%) | 210× (62–1040)[a] | 330× (73–2312)[a] |
| French gold, full exome (27; 247/270) | Flank | 40/62 (65%) | 74/75 (99%) | 133/133 (100%) | 39× (30–49) | 49× (35–62) |
| French gold, full exome (27; 247/270) | Window | 35/57 (61%) | 66/67 (99%) | 146/146 (100%) | 46× (33–59) | 58× (39–78) |
| German gold, full exome (19; 140/190) | Flank | 5/27 (19%) | 29/41 (71%) | 106/122 (87%) | 177× (95–322) | 260× (123–523)[a] |
| German gold, full exome (19; 140/190) | Window | 18/50 (36%) | 39/52 (75%) | 83/88 (94%) | 114× (63–185) | 166× (82–293)[a] |
| Gold pooled, full exome (46; 387/460) | Flank | 45/89 (51%) | 103/116 (89%) | 239/255 (94%) | 97× (56–170) | 152× (75–310) |
| Gold pooled, full exome (46; 387/460) | Window | 53/107 (50%) | 105/119 (88%) | 229/234 (98%) | 75× (50–107) | 106× (64–159) |
| German CeRKiD, slice (14; 242/392) | Flank | 31/107 (29%) | 77/121 (64%) | 134/164 (82%) | 207× (93–534)[a] | 335× (130–1054)[a] |
| German CeRKiD, slice (14; 242/392) | Window | 47/149 (32%) | 95/126 (75%) | 100/117 (85%) | 142× (67–418)[a] | 215× (88–851)[a] |
| Gold, slice, sensitivity analysis (46; 1090/1288) | Flank | 139/276 (50%) | 302/332 (91%) | 649/680 (95%) | 82× (53–131) | 119× (68–221) |
| Gold, slice, sensitivity analysis (46; 1090/1288) | Window | 169/318 (53%) | 314/352 (89%) | 607/618 (98%) | 66× (47–92) | 90× (59–132) |

| Stratum (samples; detected/runs) | Depth | <50× | 50–100× | ≥100× | Depth for 90% (95% CI) | Depth for 95% (95% CI) |
|---|---|---|---|---|---|---|
| All samples, all seeds, sensitivity analysis (60; 629/852) | Flank | 76/196 (39%) | 180/237 (76%) | 373/419 (89%) | 147× (87–247) | 235× (125–448)[a] |
| All samples, all seeds, sensitivity analysis (60; 629/852) | Window | 100/256 (39%) | 200/245 (82%) | 329/351 (94%) | 104× (66–165) | 151× (88–275) |

Detection in 600 pooled runs of 60 downsampled confirmed positive exomes (VNtyper 2.0.47): French and German gold full exomes and German CeRKiD region slices (Supplementary Methods S9). Pooled and full-depth strata rows use one run per sample and fraction; all-seeds and gold slice rows are sensitivity analyses. Columns <50× to ≥100× give detected/runs by depth band; strata by full-depth confidence tier are post hoc. Flank, mean depth over the unique VNTR flanks; Window, GRCh38 mean depth over the VNTR window used by the coverage verdict. Depth for 90% and 95%, depth at which the logistic GEE reaches that sensitivity, with 95% cluster bootstrap CI. Only unflagged calls count as detection. All samples are positive (sensitivity only).

[a] The upper confidence bound lies beyond the highest observed depth.

#### Supplementary Table S12: Paired KAPA and Twist exomes of the same carriers

| Sample | PacBio | Kit | VNtyper 2 result | Flank depth, QC | VNTR reads | Bases < Q20 (%) | Clean reads (%) |
|---|---|---|---|---|---|---|---|
| CZUS-015 | 23dupC | KAPA HEv2 | HP call of a different event (C>CGGCT), not the PacBio variant | 266×, PASS | 1888 | 21.7 | 14 |
| CZUS-015 | 23dupC | Twist v2 | HP +1 frameshift, 41 alt reads, score 0.00725 | 133×, REDUCED | 1940 | 10.1 | 56 |
| CZUS-004 | 54_55insG | KAPA HEv2 | negative | 278×, PASS | 2347 | 21.3 | 14 |
| CZUS-004 | 54_55insG | Twist v2 | HP +1 frameshift, 48 alt reads, score 0.00675 | 143×, PASS | 2576 | 9.6 | 58 |
| CZUS-009 | 58_59insG | KAPA HEv2 | negative | 172×, PASS | 2977 | 22.8 | 11 |
| CZUS-009 | 58_59insG | Twist v2 | below threshold (0.00467) | 150×, PASS | 4203 | 11.3 | 52 |

| Sample | PacBio | Kit | VNtyper 2 result | Flank depth, QC | VNTR reads | Bases < Q20 (%) | Clean reads (%) |
|---|---|---|---|---|---|---|---|
| Czech screening | 59dupC | KAPA HEv2 | LP +1 frameshift, 10 alt reads, score 0.00588 | 101×, FAIL | 771 | 11.0 | 47 |
| Czech screening | 59dupC | Twist v2 | HP +1 frameshift, 76 alt reads, score 0.01377 | 131×, PASS | 2245 | 10.9 | 52 |

Four carriers missed on KAPA HyperExome V2 (HEv2) and resequenced from the same DNA with Twist Exome v2 (Supplementary Methods S11), analysed with VNtyper 2.0.47. Result, unflagged Kestrel call (HP, high precision; LP, low precision) or, for negatives, the highest depth score below the reporting threshold; flank depth, mean depth over the unique VNTR flanks; QC, coverage verdict; VNTR reads, non-duplicate reads carrying repeat sequence; bases < Q20, share of their bases below base quality 20; clean reads, share of VNTR reads with fewer than 5% such bases. KAPA results come from this rerun of the transferred exomes and differ slightly from Table S8, which reports the contributing centre's run: flank depths by a few reads, and for CZUS-015 an unflagged call of C>CGGCT, not the PacBio variant, where the centre's run was negative. C>CGGCT is a recurrent candidate in KAPA exomes, present below the threshold in 9 of 263 Czech samples, and the counterpart of the filtered C>CGGCA artifact; small input differences decide whether Kestrel's haplotype search reaches it.

### Supplementary Data Files

Per-sample tables are on Zenodo as **Data S1-S4**. Data S1, preregistered simulation results for the natural-length and long-allele simulations: all per-sample results, and performance by arm, variant type, coverage fraction, allele length and method combination. Data S2, de-identified validation and screening calls (final unflagged, candidate and flagged). Data S3, reference method, result and variant of each reference-standard sample. Data S4, one row per reference-standard sample with cohort, capture kit, assembly, VNTR flank and mean depth, coverage QC verdict, reference result, and the VNtyper 1.3.0 and VNtyper 2.0.47 calls (Supplementary Methods S7, S10). DOI: 10.5281/zenodo.20014840 (concept DOI; current version: 10.5281/zenodo.20015364).

### ADTKD-Net Consortium Acknowledgements

We thank the following ADTKD-Net consortium members for their contributions to the consortium discussions and consensus that informed the proposed testing workflow. These individuals are not listed as authors on this manuscript and the acknowledgement here does not imply ICMJE authorship.

- Roser Torra, Nephrology Department, Fundació Puigvert, Institut de Recerca Sant Pau, Universitat Autònoma de Barcelona, Barcelona, Spain (ORCID: 0000-0001-8714-2332).
- Olivier Devuyst, Institute of Physiology, University of Zurich, Zurich, Switzerland (ORCID: 0000-0003-3744-4767).
- Eric Olinger, Center for Human Genetics, Cliniques universitaires Saint-Luc, UCLouvain, Brussels, Belgium (ORCID: 0000-0003-1178-7980).
- Ewout J. Hoorn, Division of Nephrology and Transplantation, Department of Internal Medicine, Erasmus MC, University Medical Center Rotterdam, Rotterdam, The Netherlands (ORCID: 0000-0002-8738-3571).
- Kerstin Amann, Department of Nephropathology, University Hospital Erlangen, Friedrich-Alexander-Universität Erlangen-Nürnberg, Erlangen, Germany (ORCID: 0000-0001-6116-8315).
- Steven P. Sourbron, School of Medicine and Population Health, University of Sheffield, Sheffield, United Kingdom (ORCID: 0000-0002-3374-3973).
- Luca Rampoldi, Molecular Genetics of Renal Disorders Unit, Division of Genetics and Cell Biology, IRCCS San Raffaele Scientific Institute, Milan, Italy (ORCID: 0000-0002-0544-7042).
- Dominik Seelow, Exploratory Diagnostic Sciences, Berlin Institute of Health at Charité – Universitätsmedizin Berlin, Berlin, Germany (ORCID: 0000-0002-9746-4412).
- Franz Schaefer, Division of Pediatric Nephrology, Center for Pediatrics and Adolescent Medicine, Heidelberg University Hospital, Heidelberg, Germany (ORCID: 0000-0001-7564-9937).
- José Antonio Ramírez García, Division of Pediatric Nephrology, Center for Pediatrics and Adolescent Medicine, Heidelberg University Hospital, Heidelberg, Germany.
- Gregory Papagregoriou, Molecular Medicine Research Center and biobank.cy Center of Excellence in Biobanking and Biomedical Research, University of Cyprus, Nicosia, Cyprus (ORCID: 0000-0002-2440-9789).

We also thank Christian Scheidler (ePAG patient representative, ADTKD-Net / European Reference Network for Rare Kidney Diseases (ERKNet)) for representing the patient perspective in ADTKD-Net discussions.

### References

S1. Audano PA, Ravishankar S, Vannberg FO. Mapping-free variant calling using haplotype reconstruction from k-mer frequencies. Berger B, ed. Bioinformatics. 2018;34(10):1659-1665. doi:10.1093/bioinformatics/btx753

S2. Saei H, Morinière V, Heidet L, et al. VNtyper enables accurate alignment-free genotyping of MUC1 coding VNTR using short-read sequencing data in autosomal dominant tubulointerstitial kidney disease. iScience. 2023;26(7):107171. doi:10.1016/j.isci.2023.107171

S3. Park J, Bakhtiari M, Popp B, et al. Detecting tandem repeat variants in coding regions using code-adVNTR. iScience. 2022;25(8):104785. doi:10.1016/j.isci.2022.104785

S4. Ji D, Aboukhalil R, Moshiri N. ViralWasm: A client-side user-friendly web application suite for viral genomics. Birol I, ed. Bioinformatics. 2024;40(1):btae018. doi:10.1093/bioinformatics/btae018

S5. Popp B, Saei H. MucOneUp: A Simulation Framework for MUC1-VNTR Variant Benchmarking. Published online May 2026. doi:10.64898/2026.05.08.723876

S6. Živná M, Kidd K, Přistoupilová A, et al. Noninvasive Immunohistochemical Diagnosis and Novel MUC1 Mutations Causing Autosomal Dominant Tubulointerstitial Kidney Disease. Journal of the American Society of Nephrology: JASN. 2018;29(9):2418-2431. doi:10.1681/ASN.2018020180

S7. Vrbacká A, Přistoupilová A, Kidd KO, et al. Single-Molecule Real-Time Sequencing for MUC1 Variable Number of Tandem Repeat Variation to Improve Autosomal Dominant Tubulointerstitial Kidney Disease Diagnosis. Journal of the American Society of Nephrology. 2026;37(10):2248-2261. doi:10.1681/ASN.0000001103

S8. Wilson EB. Probable Inference, the Law of Succession, and Statistical Inference. Journal of the American Statistical Association. 1927;22(158):209-212. doi:10.1080/01621459.1927.10502953

S9. Newcombe RG. Two-sided confidence intervals for the single proportion: Comparison of seven methods. Statistics in Medicine. 1998;17(8):857-872. doi:10.1002/(sici)1097-0258(19980430)17:8<857::aid-sim777>3.0.co;2-e

S10. Ekici AB, Hackenbeck T, Morinière V, et al. Renal fibrosis is the common feature of autosomal dominant tubulointerstitial kidney diseases caused by mutations in mucin 1 or uromodulin. Kidney International. 2014;86(3):589-599. doi:10.1038/ki.2014.72

S11. Wenzel A, Altmueller J, Ekici AB, et al. Single molecule real time sequencing in ADTKD-MUC1 allows complete assembly of the VNTR and exact positioning of causative mutations. Scientific Reports. 2018;8(1):4170. doi:10.1038/s41598-018-22428-0

S12. Kachmar J, Saei H, Morinière V, et al. Phenotypic Heterogeneity of ADTKD-MUC1 Diagnosed Using VNtyper, a Novel Genetic Technique. American Journal of Kidney Diseases: The Official Journal of the National Kidney Foundation. 2025;85(5):603-609.e1. doi:10.1053/j.ajkd.2024.11.010

S13. World Medical Association. World Medical Association Declaration of Helsinki: Ethical principles for medical research involving human subjects. JAMA. 2013;310(20):2191-2194. doi:10.1001/jama.2013.281053

S14. Kirby A, Gnirke A, Jaffe DB, et al. Mutations causing medullary cystic kidney disease type 1 lie in a large VNTR in MUC1 missed by massively parallel sequencing. Nature Genetics. 2013;45(3):299-303. doi:10.1038/ng.2543

S15. DerSimonian R, Laird N. Meta-analysis in clinical trials. Controlled Clinical Trials. 1986;7(3):177-188. doi:10.1016/0197-2456(86)90046-2

S16. IntHout J, Ioannidis JPA, Borm GF. The Hartung-Knapp-Sidik-Jonkman method for random effects meta-analysis is straightforward and considerably outperforms the standard DerSimonian-Laird method. BMC medical research methodology. 2014;14:25. doi:10.1186/1471-2288-14-25

S17. Röver C, Knapp G, Friede T. Hartung-Knapp-Sidik-Jonkman approach and its modification for random-effects meta-analysis with few studies. BMC medical research methodology. 2015;15:99. doi:10.1186/s12874-015-0091-1

S18. Denti L, Pirola Y, Previtali M, et al. Shark: Fishing relevant reads in an RNA-Seq sample. Ponty Y, ed. Bioinformatics. 2021;37(4):464-472. doi:10.1093/bioinformatics/btaa779

S19. Bensouna I, Robert T, Vanhoye X, et al. Systematic Screening of Autosomal Dominant Tubulointerstitial Kidney Disease–MUC1 27dupC Pathogenic Variant through Exome Sequencing. Journal of the American Society of Nephrology. 2025;36(2):256-263. doi:10.1681/ASN.0000000503
